# Timing and Dose of Pharmacological Thromboprophylaxis in Adult Trauma Patients: Perceptions, Barriers, and Experience of Saudi Arabia Practicing Physicians

**DOI:** 10.1101/2021.01.26.21250366

**Authors:** Marwa Amer, Mohammed Bawazeer, Khalid Maghrabi, Rashid Amin, Edward De Vol, Mohammed Hijazi

**Affiliations:** Pharmaceutical Care Division, King Faisal Specialist Hospital and Research Center, Riyadh, Saudi Arabia; College of Medicine, Alfaisal University; Department of Critical Care Medicine, King Faisal Specialist Hospital and Research Center, Riyadh, Saudi Arabia; Department of Biostatistics, Epidemiology, and Scientific Computing, King Faisal Specialist Hospital and Research Center, Riyadh, Saudi Arabia

**Author notes:** **Corresponding Authors: Marwa Amer PharmD, B.S. Pharm, BCPS, BCCCP**. Both contributed equally as first Author (Marwa Amer, Mohammed Bawazeer). **Authors contribution** MA, MB: conception and design, analytical plan, drafting of the manuscript, critical revision of the manuscript for important intellectual content, approval of the final version to be published and both contributed equally as first Author. KM, RA, MH: acquisition, analysis or interpretation of data, critical revision of the manuscript for important intellectual content, and approval of the final version to be published. ED: performed statistical analysis, revision of the manuscript. **Funding:** None. Marwa Amer PharmD, BCPS, BCCCP Critical Care Pharmacy Specialist, Medical and Surgical ICU Adjunct Assistant Professor- Alfaisal University- College Medicine Pharmaceutical Care Division (MBC # 11) King Faisal Specialist Hospital & Research Center PO Box 3354, Riyadh, 11211. Mohammed Bawazeer MD, FRCSC, FACS Trauma & Acute Care Surgery Consultant Intensivist Medical Director, Surgical Critical Care Unit Founder and Head of the Surgical Critical Care Chapter, Saudi Critical Care Society Department of Critical Care Medicine (MBC 94) King Faisal Specialist Hospital & Research Centre PO Box 3354, Riyadh 11211. Khalid Maghrabi, MD, FCCP, MRCP (UK) Consultant Intensivist Department of Critical Care Medicine (MBC 94) King Faisal Specialist Hospital & Research Centre PO Box 3354, Riyadh 11211. Rashid Amin PharmD, BCPS, BCCCP Critical Care Pharmacy Specialist, Medical and Surgical ICU Pharmaceutical Care Division (MBC # 11) King Faisal Specialist Hospital & Research Center PO Box 3354, Riyadh, 11211. Edward De Vol Chairman Department of Biostatistics, Epidemiology, and Scientific Computing King Faisal Specialist Hospital & Research Centre PO Box 3354, Riyadh 11211. Mohammed Hijazi, MD, FACP, FCCP Consultant Intensivist Department of Critical Care Medicine (MBC 94) King Faisal Specialist Hospital & Research Centre PO Box 3354, Riyadh 11211.

**Keywords:** Venous thromboembolism (VTE), pharmacological venous thromboembolism prophylaxis (PVTE-Px), augmented renal clearance (ARC), traumatic brain injury (TBI), spinal cord injury (SCI), non-operative (NOR) solid organ injuries, conservatively managed solid organ injuries, low molecular weight heparin (LMWH), unfractionated heparin (UFH), adult trauma patients

## Abstract

**Background:** Pharmacological venous thromboembolism prophylaxis (PVTE-Px) in trauma care is challenging and frequently delayed until post injury bleeding risk is perceived to be sufficiently low; yet data for optimal initiation time is lacking. This study assessed practice pattern of PVTE-Px initiation time and dose in traumatic brain injury (TBI), spinal cord injury (SCI), and non-operative (NOR) solid organ injuries.

**Methods:** Multicenter, cross sectional, observational, survey-based study involving intensivists, trauma surgeons, general surgeons, spine orthopedics, and neurosurgeons practicing in trauma centers. The data of demographics, PVTE-Px timing and dose, and five clinical case scenarios were obtained. Analyses were stratified by early initiators vs. late initiators and logistic regression models were used to identify factors associated with early initiation of PVTE-Px.

**Results:** Of 102 physicians (29 % response rate), most respondents were intensivists (63.7%) and surgeons (who are general and trauma surgeons) (22.5%); majority were consultants (58%), practicing at level 1 trauma centers (40.6%) or academic teaching hospitals (45.1%). A third of respondents (34.2%) indicated that decision to initiate PVTE-Px in TBI and SCI was made by a consensus between surgical, critical care, and neurosurgical services. For patients with NOR solid organ injuries, 34.2% of respondents indicated trauma surgeons initiated the decision on PVTE-Px timing. About 53.7% of the respondents considered their PVTE-Px practice as appropriate, half used combined mechanical and PVTE-Px (57.1%), 52% preferred enoxaparin (40 mg once daily), and only 6.5% used anti-Xa level to guide enoxaparin prophylactic dose. Responses to clinical cases varied. For TBI and TBI with intracranial pressure monitor, 40.3% and 45.6% of the respondents were early initiators with stable repeated head computed tomography [CT], respectively. For SCI, most respondents were early initiators without repeated CT spine (36.8%). With regards to NOR solid organ injuries [gunshot wound to the liver and grade IV splenic injuries], 49.1% and 36.4% of respondents were early initiators without a repeat CT abdomen.

**Conclusions:** Variations were observed in PVTE-Px initiation time influenced by trauma type. Our findings suggested enoxaparin is preferred in a standard prophylactic dose. More robust data from randomized trials are needed and the use of clinicians’ judgment is recommended.

**Key Messages:**

1. Ideal time to initiate therapy, agent selection, dosing, and monitoring of pharmacological venous thromboembolism prophylaxis (PVTE-Px) for trauma patients is challenging.
2. Variations were observed in PVTE-Px initiation time influenced by trauma type.
3. Our study results are relatively in line with the recent evidence-based clinical literature
4. Our findings suggested limited awareness of augmented renal clearance (ARC) and utilization of serum anti-factor-Xa (anti-Xa) level.

## Introduction

Venous thromboembolism (VTE) is a potentially life-threatening complication that can develop after traumatic injury with a reported incidence of deep venous thrombosis (DVT) of up to 63% and pulmonary embolism (PE) of up to 22% [1,2]. Moreover, the current estimates of DVT and PE are around 5% and 2%, respectively, for adult patients with severe injury who are receiving pharmacological venous thromboembolism prophylaxis (PVTE-Px) [3,4]. The Eastern Association for the Surgery of Trauma (EAST) and the American College of Chest Physicians (ACCP) have developed and published guidelines for PVTE-Px in 2002 and 2012, respectively [5,6]. Both guidelines recognize the importance of initiating PVTE-Px, but there is limited evidence to date to conclude robust recommendations regarding the timing and dose of PVTE-Px. A recent Cochrane review by Barrera *et al* concluded that any form of thromboprophylaxis (mechanical and/or pharmacological) reduced the risk of DVT, but did not affect mortality or rate of PE [7]. Specifically, the use of PVTE-Px was more effective than mechanical devices at reducing the risk of DVT. The use of low molecular weight heparin (LMWH) appeared to decrease the risk of DVT when compared with unfractionated heparin (UFH) for thromboprophylaxis. The authors recommended PVTE-Px in trauma patients, but again this lacked specific recommendations regarding timing, and dose. Of note, we will be using pharmacological thromboprophylaxis interchangeably with PVTE-Px throughout the entire manuscript.

Traumatic brain injury (TBI) represents a major cause of death and disability as demonstrated by the 2.5 million emergency department (ED) visits, 282,000 hospitalizations, and 56,000 deaths in 2013 [8]. Although thus far most trauma surgeons agree that PVTE-Px is necessary in TBI patients, it is frequently delayed until the risk of post injury bleeding is perceived to be sufficiently low. Despite the general coherency of this rationale, there is considerable uncertainty about the optimal timing of safe initiation of PVTE-Px. In addition, there is variation in clinical practice guideline recommendations regarding this topic and a review has been previously published [9,10,11,12,13,14], **[supplementary table 1]**. A survey of EAST members revealed significant variation in practice in terms of initiating PVTE-Px for TBI patients [15]. Additionally, there is also no consensus regarding safety of initiating PVTE-Px in the presence of intracranial pressure (ICP) monitors, or post craniotomy. Those patients are categorized as high risk in American College of Surgeons, Trauma Quality Improvement Program (ACS-TQIP) which suggested inferior vena cava filter placement for VTE prevention [13]. One retrospective review found no association of increased hemorrhage with the use of LMWH with invasive ICP monitors. The median length of time to a stable head computed tomography (CT) in this study was 2 days, and the median time to initiation of PVTE-Px was 3.6 days [16].

**Table 1:** Demographics, practitioner and practice site baseline characteristics Table legend: $ Surgeons include trauma surgeons and general surgeons * Other specialties include orthopedic surgeons and neurosurgeons # Other (free text): Blast injuries, gunshot injuries, war injuries, burns, industrial injuries, cervical spine injuries, internal bleeding, and polytrauma Abbreviations: ICU: intensive care unit

|  | Total cohort<br>N/# of responses<br>(%) | Intensivists<br>N/# of<br>responses (%) | Surgeons §<br>N/# of<br>responses (%) | Other specialties *<br>N/# of responses (%) | P value |
| --- | --- | --- | --- | --- | --- |
| **Position** |  |  |  |  |  |
| Fellow | 24/100 (24) | 19 / 64 (29.69) | 2/ 22 (9.09) | 3/14 (21.43) | 0.1820 |
| Associate consultant | 18/100 (18) | 11/64 (17.29) | 3/ 22 (13.64) | 4/ 14 (28.57) |  |
| Consultant | 58/100 (58) | 34/64 (53.13) | 17/22 (77.27) | 7 /14 (50) |  |
| **Practicing years** |  |  |  |  |  |
| <5 years | 47/101(46.5) | 31/ 65 (47.69) | 10/22 (45.45) | 6/14 (42.86) | 0.9210 |
| 5-10 years | 21/101 (20.79) | 14/65 (21.54) | 5/22 (22.73) | 2/14 (14.29) |  |
| > 10 years | 33/101 (32.67) | 20 / 65 (30.77) | 7/22 (31.82) | 6/14 (42.86) |  |
| **Certification board of your primary specialty** |  |  |  |  |  |
| <10 years | 40/90 (44.44) | 25/57 (43.86) | 9 /21 (42.86) | 6/12 (50) | 0.3680 |
| 10-20 years | 40/90 (44.44) | 28 /57 (49.12) | 9 /21 (42.86) | 3/12 (25) |  |
| >20 years | 10/90 (11.11) | 4 /57 (7.02) | 3/ 21 (14.29) | 3/12 (25) |  |
| **Geographic location (geographic)** |  |  |  |  |  |
| Central region | 62/99 (62.63) | 39/64 (60.94) | 18/22 (81.82) | 5/13 (38.46) | 0.0275 |
| Northern region | 6/99 (6.06) | 4/64 (6.25) | 0 | 2/13 (15.38) |  |
| Western region | 13/99 (13.13) | 12/64 (18.75) | 0 | 1/13 (7.69) |  |
| Eastern region | 11/99 (11.11) | 6/64 (9.38) | 2/22 (9.09) | 3/13 (23.08) |  |
| Southern region | 7/99 (7.07) | 3/64 (4.69) | 2/22 (9.09) | 2/13 (15.38) |  |
| **Trauma center** |  |  |  |  |  |
| Level 1 trauma center | 41/101 (40.59) | 22/65 (33.85) | 13/22 (59.09) | 6/14 (42.86) | 0.1040 |
| Level 2 trauma center | 24/101 (23.76) | 16/65 (24.62) | 5/22 (22.73) | 3/14 (21.43) |  |
| Level 3 trauma center | 9/101 (8.91) | 6/65 (9.23) | 0 | 3/14 (21.43) |  |
| Undesignated | 27/101 (26.73) | 21/65 (32.31) | 4/22 (18.18) | 2/14 (14.29) |  |
| **No. of trauma cases seen per year** |  |  |  |  |  |
| <50 cases | 42/101 (41.58) | 36/65 (55.38) | 4/22 (18.18) | 2/14 (14.29) | 0.0009 |
| 50-100 cases | 21/101 (20.79) | 13/65 (20) | 6/22 (27.27) | 2/14 (14.29) |  |
| >100 cases | 38/ 101 (37.62) | 16/65 (24.62) | 12/22 (54.55) | 10/14 (71.43) |  |
| **No. of ICU beds in your facility** |  |  |  |  |  |
| <20 beds | 16/101 (15.84) | 8/65 (12.31) | 3/22 (13.64) | 5/14 (35.71) | 0.3965 |
| 20-60 beds | 59/101 (58.42) | 40/65 (61.54) | 13/22 (59.09) | 6/14 (42.86) |  |
| >60 beds | 26/101 (25.74) | 17/65 (26.15) | 6/22 (27.27) | 3/14 (21.43) |  |
| **Practice type (select all that apply )** |  |  |  |  |  |
| Academic teaching hospital | 46/102 (45.1) | 30/65 (46.15) | 14/ 23 (30.43) | 2/14 (14.29) | 0.0148 |
| Ministry of health hospital | 36/ 102 (35.29) | 20/65 (30.77) | 8/23 (34.78) | 8/14 (57.14) | 0.1860 |
| Private hospital | 10/102 (9.8) | 7/65 (10.77) | 2/23 (8.7) | 1/14 (7.14) | 0.8948 |
| Government (national guard, military) | 28/102 (27.45) | 19/65 (29.23) | 6/23 (26.09) | 3/14 (21.43) | 0.822 |

| Type of trauma seen (select all that apply ) | Total cohort<br>N/# of responses (%) | Intensivists<br>N/# of responses (%) | Surgeons<sup>\$</sup><br>N/# of responses (%) | Other specialties *<br>N/# of responses (%) | P value |
| --- | --- | --- | --- | --- | --- |
| Fall | 63/102 (61.76) | 40/65 (61.54) | 13/23 (56.52) | 10/14 (71.43) | 0.6565 |
| Motor vehicle accidents | 89/102 (87.25) | 55/65 (84.62) | 20/23 (86.96) | 14/14 (100) | 0.1222 |
| Motorcycle | 38/102 (37.25) | 22/65 (33.85) | 9/23 (39.13) | 7/14 (50) | 0.5214 |
| Pedestrian | 53 / 102 (51.96) | 33/65 (50.77) | 12/23 (52.17) | 8/14 (57.14) | 0.91 |
| Blunt | 45/102 (44.12) | 24/65 (36.92) | 14/23 (60.87) | 7/14 (50) | 0.1238 |
| Stab | 35/102 (34.31) | 21/65 (32.31) | 8/23 (34.78) | 6/14 (42.86) | 0.7565 |
| Intracranial injuries | 60/102 (58.82) | 38/65 (58.46) | 12/23 (52.17) | 10/14 (71.43) | 0.5022 |
| Cerebral contusion or laceration | 49/102 (48.04) | 28/65 (43.08) | 12/23 (52.17) | 9/14 (64.29) | 0.3173 |
| Brainstem or cerebellar injury | 37/102 (36.27) | 21/65 (32.31) | 8/23 (34.78) | 8/14 (57.14) | 0.2250 |
| Pelvic fracture | 52/102 (50.98) | 31/65 (47.69) | 13/23 (56.52) | 8/14 (57.14) | 0.6776 |
| Femur shaft fracture | 48/102 (47.06) | 30/65 (46.15) | 12/23 (52.17) | 6/14 (42.86) | 0.8344 |
| Tibia or fibula fracture | 44/102 (43.14) | 27/65 (41.54) | 11/23 (47.83) | 6/14 (42.86) | 0.8724 |
| Spinal cord injury | 25/102 (24.51) | 15/65 (23.08) | 5/23 (21.74) | 5/14 (35.71) | 0.5934 |
| Other (free text) <sup>#</sup> | 4/102 (3.92) | 2/65 (3.08) | 2/23 (8.7) | 0 | 0.3170 |

Patients with a spinal cord injury (SCI) represent a subset of trauma patients that have a substantial risk of developing VTE complications with an incidence of DVT and PE ranging from 49% to 72% [17]. While it makes intuitive sense that the early initiation of PVTE-Px would lower the incidence of VTE events, it can also result in some serious hemorrhagic complications, including spinal hematoma. On the other hand, Tracy *et al* evaluated the effects of delayed prophylaxis in patients with TBI and SCI. The authors concluded that patients who developed thrombotic complications had significantly longer times (6.7 ± 4.9 d) until the initiation of PVTE-Px. For each day there was a delay in time to initiation, the odds of VTE significantly increased [18]. Similar to TBI, the timing of PVTE-Px initiation is not well-defined in the guidelines and remains controversial [9,11,12,19].

Injuries to solid organs such as the spleen, kidney, and liver are frequently encountered in patients with abdominal trauma [20]. The management of solid-organ injuries has evolved from a primarily operative approach to a predominantly nonoperative approach (i.e., observation and serial hemoglobin monitoring or angioembolization for site of bleeding) in the hemodynamically stable patient [21,22]. With management shifting from an operative to an observational approach, clinicians must be cognizant that the initiation of PVTE-Px may have to be delayed for at least 24 hours following the stabilization of hemoglobin [23]. The EAST practice management guidelines for blunt hepatic and splenic injury stated that PVTE-Px could be used, yet there is limited evidence to date to conclude robust recommendations regarding the timing and dose of PVTE-Px [9, 24,25].

Regarding the specific PVTE-Px agent for the trauma patients, recent literature suggests that LMWH use is associated with a lower rate of DVT and PE than UFH [26]. Therefore, the 2002 EAST Guidelines recommend LMWH use for VTE prophylaxis in trauma patients with certain injuries (pelvic fractures, complex lower-extremity fractures, and SCI) if they are not at a high risk of bleeding (level II recommendation) and as the primary method of VTE prophylaxis in trauma patients with an Injury Severity Score (ISS) >9 (level III recommendation, low-quality evidence) [5]. On the other hand, practitioners might favor the shorter half-life of UFH in patients where the perceived risk of hemorrhagic complications is high [27]. Regarding the dose, neither the ACCP nor the EAST practice management guidelines give a recommendation of LMWH specific thromboprophylaxis dosing. Trauma, TBI, and SCI patients are at risk of augmented renal clearance (ARC) which is reported to be 67-85 % in those population resulting in subtherapeutic drug concentration [28]. Recent studies have shown that subtherapeutic anti-Xa levels are more frequent in trauma patients than originally thought [29]. Therefore, alternative regimens in the general trauma population have been suggested in the literature [30,31,32,33,34] **[supplementary table 2]**.

**Table 2:**
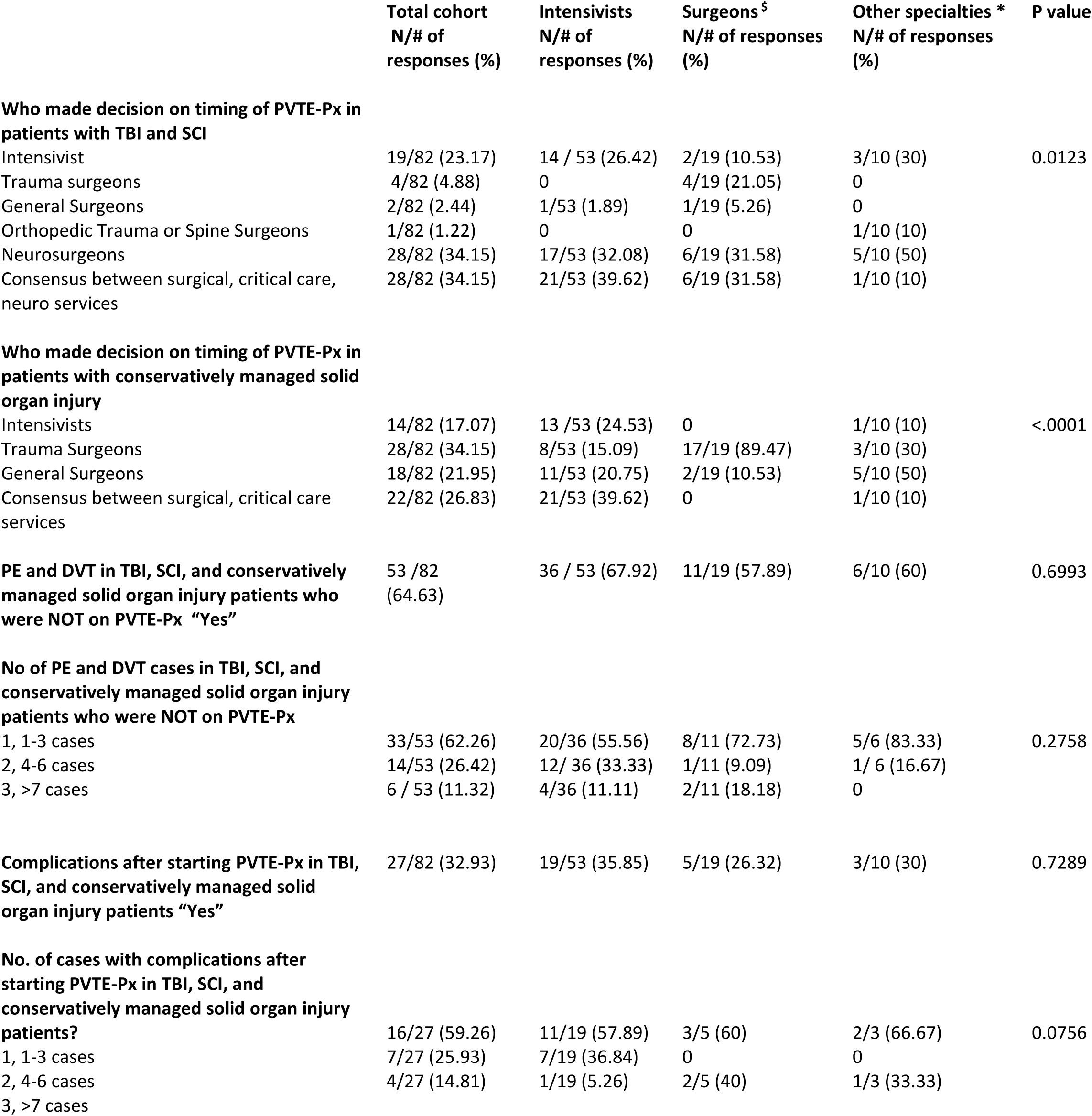
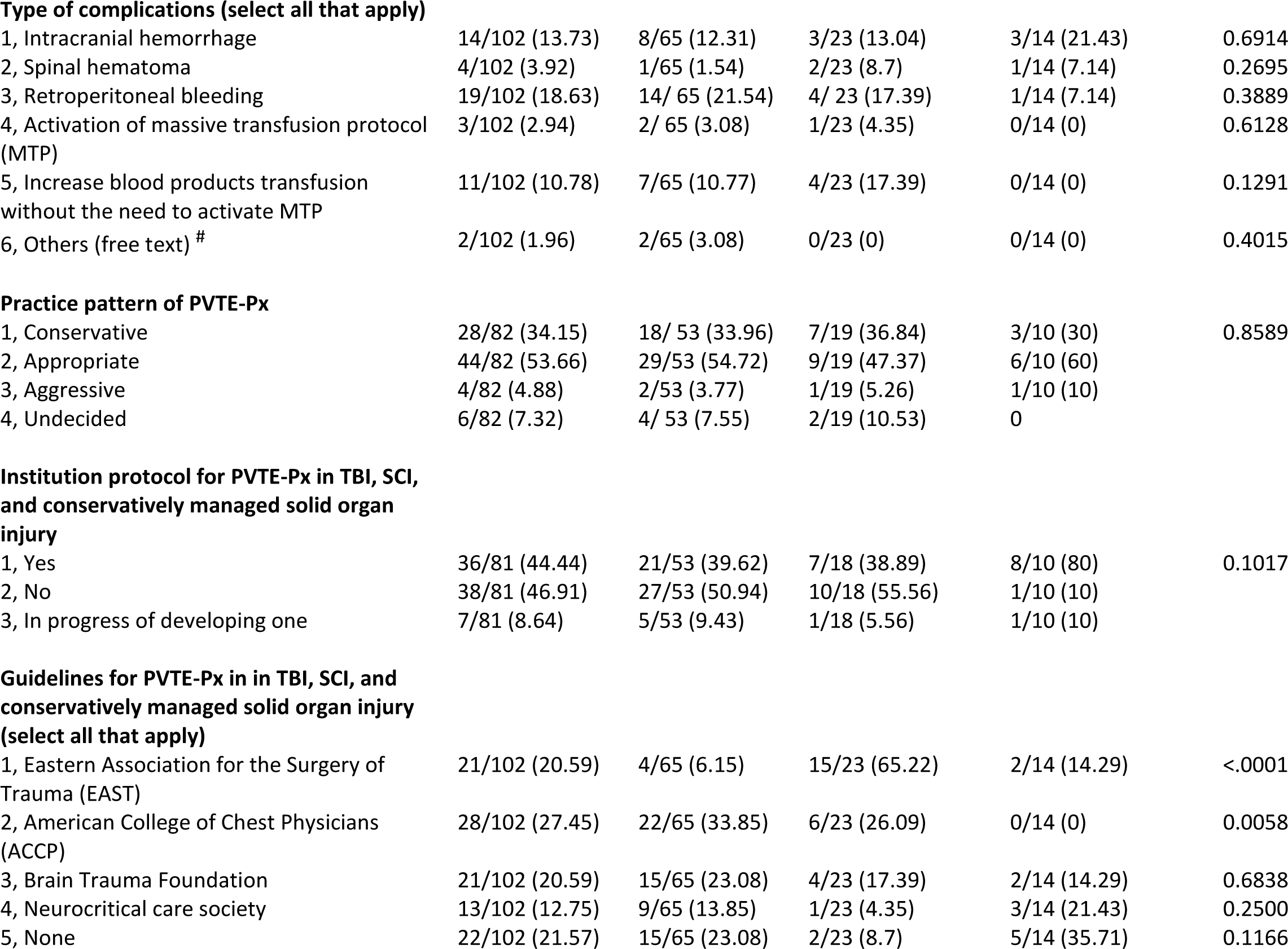
PVTE-Px general questions Table legend: $ Surgeons includes trauma surgeons, general surgeons * Other specialties includes orthopedic surgeons, neurosurgeons # Other (free text): ETT bleeding, UGIB, pulmonary hemorrhage Abbreviations: PVTE-Px: pharmacological venous thromboembolism prophylaxis, TBI: traumatic brain injury, SCI: spinal cord injury, DVT: deep vein thrombosis, ETT: endotracheal tube, UGIB: upper gastrointestinal bleeding

|  | Total cohort<br>N/# of<br>responses (%) | Intensivists<br>N/# of<br>responses (%) | Surgeons<sup>§</sup><br>N/# of responses<br>(%) | Other specialties *<br>N/# of responses<br>(%) | P value |
| --- | --- | --- | --- | --- | --- |
| **Who made decision on timing of PVTE-Px in patients with TBI and SCI** |  |  |  |  |  |
| Intensivist | 19/82 (23.17) | 14 / 53 (26.42) | 2/19 (10.53) | 3/10 (30) | 0.0123 |
| Trauma surgeons | 4/82 (4.88) | 0 | 4/19 (21.05) | 0 |  |
| General Surgeons | 2/82 (2.44) | 1/53 (1.89) | 1/19 (5.26) | 0 |  |
| Orthopedic Trauma or Spine Surgeons | 1/82 (1.22) | 0 | 0 | 1/10 (10) |  |
| Neurosurgeons | 28/82 (34.15) | 17/53 (32.08) | 6/19 (31.58) | 5/10 (50) |  |
| Consensus between surgical, critical care, neuro services | 28/82 (34.15) | 21/53 (39.62) | 6/19 (31.58) | 1/10 (10) |  |
| **Who made decision on timing of PVTE-Px in patients with conservatively managed solid organ injury** |  |  |  |  |  |
| Intensivists | 14/82 (17.07) | 13 / 53 (24.53) | 0 | 1/10 (10) | <.0001 |
| Trauma Surgeons | 28/82 (34.15) | 8/53 (15.09) | 17/19 (89.47) | 3/10 (30) |  |
| General Surgeons | 18/82 (21.95) | 11/53 (20.75) | 2/19 (10.53) | 5/10 (50) |  |
| Consensus between surgical, critical care services | 22/82 (26.83) | 21/53 (39.62) | 0 | 1/10 (10) |  |
| **PE and DVT in TBI, SCI, and conservatively managed solid organ injury patients who were NOT on PVTE-Px “Yes”** | 53 /82 (64.63) | 36 / 53 (67.92) | 11/19 (57.89) | 6/10 (60) | 0.6993 |
| **No of PE and DVT cases in TBI, SCI, and conservatively managed solid organ injury patients who were NOT on PVTE-Px** |  |  |  |  |  |
| 1, 1-3 cases | 33/53 (62.26) | 20/36 (55.56) | 8/11 (72.73) | 5/6 (83.33) | 0.2758 |
| 2, 4-6 cases | 14/53 (26.42) | 12/ 36 (33.33) | 1/11 (9.09) | 1/ 6 (16.67) |  |
| 3, >7 cases | 6 / 53 (11.32) | 4/36 (11.11) | 2/11 (18.18) | 0 |  |
| **Complications after starting PVTE-Px in TBI, SCI, and conservatively managed solid organ injury patients “Yes”** | 27/82 (32.93) | 19/53 (35.85) | 5/19 (26.32) | 3/10 (30) | 0.7289 |
| **No. of cases with complications after starting PVTE-Px in TBI, SCI, and conservatively managed solid organ injury patients?** |  |  |  |  |  |
| 1, 1-3 cases | 16/27 (59.26) | 11/19 (57.89) | 3/5 (60) | 2/3 (66.67) | 0.0756 |
| 2, 4-6 cases | 7/27 (25.93) | 7/19 (36.84) | 0 | 0 |  |
| 3, >7 cases | 4/27 (14.81) | 1/19 (5.26) | 2/5 (40) | 1/3 (33.33) |  |

|  | Total cohort<br>N/# of<br>responses (%) | Intensivists<br>N/# of<br>responses (%) | Surgeons<sup>§</sup><br>N/# of responses<br>(%) | Other specialties<sup>*</sup><br>N/# of responses<br>(%) | P value |
| --- | --- | --- | --- | --- | --- |
| **Type of complications (select all that apply)** |  |  |  |  |  |
| 1, Intracranial hemorrhage | 14/102 (13.73) | 8/65 (12.31) | 3/23 (13.04) | 3/14 (21.43) | 0.6914 |
| 2, Spinal hematoma | 4/102 (3.92) | 1/65 (1.54) | 2/23 (8.7) | 1/14 (7.14) | 0.2695 |
| 3, Retroperitoneal bleeding | 19/102 (18.63) | 14/ 65 (21.54) | 4/ 23 (17.39) | 1/14 (7.14) | 0.3889 |
| 4, Activation of massive transfusion protocol (MTP) | 3/102 (2.94) | 2/ 65 (3.08) | 1/23 (4.35) | 0/14 (0) | 0.6128 |
| 5, Increase blood products transfusion without the need to activate MTP | 11/102 (10.78) | 7/65 (10.77) | 4/23 (17.39) | 0/14 (0) | 0.1291 |
| 6, Others (free text) <sup>#</sup> | 2/102 (1.96) | 2/65 (3.08) | 0/23 (0) | 0/14 (0) | 0.4015 |
| **Practice pattern of PVTE-Px** |  |  |  |  |  |
| 1, Conservative | 28/82 (34.15) | 18/ 53 (33.96) | 7/19 (36.84) | 3/10 (30) | 0.8589 |
| 2, Appropriate | 44/82 (53.66) | 29/53 (54.72) | 9/19 (47.37) | 6/10 (60) |  |
| 3, Aggressive | 4/82 (4.88) | 2/53 (3.77) | 1/19 (5.26) | 1/10 (10) |  |
| 4, Undecided | 6/82 (7.32) | 4/ 53 (7.55) | 2/19 (10.53) | 0 |  |
| **Institution protocol for PVTE-Px in TBI, SCI, and conservatively managed solid organ injury** |  |  |  |  |  |
| 1, Yes | 36/81 (44.44) | 21/53 (39.62) | 7/18 (38.89) | 8/10 (80) | 0.1017 |
| 2, No | 38/81 (46.91) | 27/53 (50.94) | 10/18 (55.56) | 1/10 (10) |  |
| 3, In progress of developing one | 7/81 (8.64) | 5/53 (9.43) | 1/18 (5.56) | 1/10 (10) |  |
| **Guidelines for PVTE-Px in in TBI, SCI, and conservatively managed solid organ injury (select all that apply)** |  |  |  |  |  |
| 1, Eastern Association for the Surgery of Trauma (EAST) | 21/102 (20.59) | 4/65 (6.15) | 15/23 (65.22) | 2/14 (14.29) | <.0001 |
| 2, American College of Chest Physicians (ACCP) | 28/102 (27.45) | 22/65 (33.85) | 6/23 (26.09) | 0/14 (0) | 0.0058 |
| 3, Brain Trauma Foundation | 21/102 (20.59) | 15/65 (23.08) | 4/23 (17.39) | 2/14 (14.29) | 0.6838 |
| 4, Neurocritical care society | 13/102 (12.75) | 9/65 (13.85) | 1/23 (4.35) | 3/14 (21.43) | 0.2500 |
| 5, None | 22/102 (21.57) | 15/65 (23.08) | 2/23 (8.7) | 5/14 (35.71) | 0.1166 |

There are still controversies and varying consensus guidelines for PVTE-Px initiation time and dose for the injured patients and in many cases they are entirely subjective, leaving it up to physician judgment as when it might be safe to initiate PVTE-Px. We hypothesized that variation observed in PVTE-Px initiation time influenced by trauma type with a considerable gap in the initiation time and dose compared to the evidence-based clinical data and clinicians’ perception. Therefore, we conducted a survey to assess the practice patterns of PVTE-Px initiation time and dose in TBI, SCI, and non-operative (NOR) solid organ injuries among Saudi Arabia practicing physicians.

## Methods

### I. Study design

This was a multicenter, cross-sectional, descriptive, observational, survey-based study in which self-administered questionnaires were used to assess the practice variation amongst intensivists, trauma surgeons, general surgeons, spine orthopedic surgeons, and neurosurgeons practicing in Saudi Arabia regarding the PVTE-Px initiation time and dose in multiple trauma patients focusing on TBI, SCI, and NOR/ conservatively managed solid organ injuries. Some of the survey questions were obtained from previous studies with some modifications to include SCI, and conservatively managed solid organ injuries [14], **[supplementary file 1]**. Once Institutional review board (IRB) approval was obtained, the survey link was distributed through email by the perspective secretaries of each national society as described below in the inclusion criteria. The survey was also distributed through the hospital health outreach services [tele-medicine]. The introduction of the survey contained the purpose of the study, and whom to contact for questions.

We used rigorous survey methodology to develop, test, and administer our questionnaire [35]. The questionnaire was mainly composed of multiple-choice questions with some open-ended questions. The survey was validated by trauma care practitioner, 3 intensivists, and 2 critical care clinical pharmacy specialists to ensure question clarity, adequacy of design, and the time needed to complete the survey. No incentives were offered for participation in the survey and data were deidentified to hospital affiliation. After initial invitation for study participation, the survey remained open for 4 months and was then re-distributed weekly. Respondents could select multiple answers (for select all that apply questions). Some of the respondents did not complete the entire survey; therefore, there were some unanswered questions. Partially completed surveys were included if at least one clinical scenario was completed. The introduction part of the survey explained the purpose of the study, confidentiality statement, and whom to contact for questions. Participants were asked at the beginning of the survey to acknowledge reading this information and voluntarily agree to participate in this research, with the knowledge that participants are free to withdraw their participation at any time. The questionnaire took approximately 10 minutes to complete. Respondents could complete the survey only once. The study was designed as an electronic survey and study data were collected and managed using institutional Research Electronic Data Capture (REDCap). The access to the RedCap data was limited to the principal investigators and co-investigators. All information was recorded anonymously and no personal identifiable information was mandatory from the participants in the survey and no patients’ related information were required in the survey. No intervention was made and there were no expected risks or direct benefits for participating in this study. The study is reported in accordance with the STrengthening the Reporting of OBservational studies in Epidemiology (STROBE) [36].

### II. Inclusion criteria

#### A. The following physicians’ specialties were included

1. Intensivists/ critical care physicians
2. Trauma surgeons
3. General surgeons
4. Spine orthopedic surgeons and
5. Neurosurgeons.

These were current member(s) of Saudi Critical Care Society, Saudi Surgical Critical Care Chapter, Saudi General Surgery Society, Spine Chapter of Saudi Orthopedic Association, and Saudi Association of Neurological Surgery practicing at level I, II, and III Trauma centers in different regions of Saudi Arabia.

#### B. Survey questions focused on

1. Traumatic brain injury (TBI)
2. Spinal cord injury (SCI), and
3. Non-operative (NOR)/ conservatively managed solid organ injuries.

### III. Exclusion

- Physicians’ specialties that were not intensivists, trauma surgeons, general surgeons, spine orthopedic surgeons, and neurosurgeons.

## End points

### Primary

- To assess the practice variation for the PVTE-Px initiation time and dose in TBI, SCI, and NOR / conservatively managed solid organ injuries amongst Saudi Arabia practicing physicians.

### Secondary

- To describe the predictors, injury characteristics, and perceived barriers for early initiations of PVTE-Px in trauma patients focusing on TBI, SCI, and NOR / conservatively managed solid organ injuries.

## Statistical analysis

Descriptive statistics were used for data analysis. Data was assumed to be non-parametric and, as such, was presented as numbers and percentages for categorical variables. Categorical data were analyzed by Fisher’s exact test or Chi-Squared test. The target population is estimated to be around 300. We hypothesized that 50% will choose to start the PVTE-Px early, allowed for 5% margin of error and 95% confidence interval (CI). Hence the required sample size was 169. Analyses were stratified by physicians’ specialties; intensivists, surgeons [including general surgeons, and trauma surgeons], and other specialties [including orthopedic surgeons, and neurosurgeons]. Analyses were further stratified by early initiator ≤ 24hr vs. late initiator >24hr in clinical scenarios related to neurotrauma (TBI, SCI), and early initiator ≤ 48hr vs. late initiator >48hr in clinical scenarios related to conservatively managed solid organ injuries. Kruskal-Wallis tests were employed for initial comparison among three or more groups. Logistic regression models were constructed to identify predictors associated with early initiations of PVTE-Px according to the injury characteristics and the perceived barriers for each clinical scenario involved. Variables included in logistic regression analysis were chosen after they were determined to have a plausible effect on the outcome of interest by consensus of the investigators. Bonferroni correction was used for adjustment in logistic regression models. The statistical analyses were performed using SAS/JMP, version 14.1 (SAS Institute Inc., Cary, NC, USA) and significance was evaluated with a 0.05 threshold using two-tailed tests of hypotheses.

## Results

The survey was sent to 350 physicians, and the response rate was 29 % (102) **[Table 1 demographics, practitioner and practice site baseline characteristics]**. The respondents were 65/102 (63.7 %) intensivists, 23/102 (22.5 %) surgeons (who are general and trauma surgeons), and 14/102 (13.7%) other specialties (who are neurosurgeons and orthopedic surgeons). The majority were consultants (58%), practicing at level 1 trauma centers (40.6%), and at academic teaching hospitals (45.1%). Among the respondents, 44.4 % were board certified of their primary specialties in 10-20 years. The major types of trauma reported were falls 61.8%, motor vehicle accidents [MVA] (87.3%), intracranial injuries (59.0%), and pedestrians (52%).

A third of respondents (34.2 %) indicated that the decision to initiate PVTE-Px in TBI and SCI was made by multidisciplinary consensus between surgical, critical care, and neurosurgical services. For patients with NOR solid organ injuries, 34.2 % of the respondents indicated that trauma surgeons usually initiate the decision on PVTE-Px timing **[Table 2: PVTE-Px general questions]**. While 64.6 % of respondents reported experience of seeing VTE without PVTE-Px, about 32.9 % witnessed complications after early PVTE-Px. The most common observed complications after early initiation of PVTE-Px were retroperitoneal bleeding (18.6 %), intracranial hemorrhage (ICH) (13.7 %), and increased blood products transfusion (10.8 %). About 53.7 % of the respondents considered their PVTE-Px practice as appropriate, 46.9 % reported absence of standardized protocol at their institution, and 21.6 % believed there were no international guidelines on early PVTE-Px initiation. Half of the respondents used combined mechanical and PVTE-Px (57.1%), 52 % preferred enoxaparin at a standard dose of 40 mg once daily, and only 6.5 % used anti-Xa level to guide enoxaparin prophylactic dose **[Table 3: preferred dose and agent selection]**.

**Table 3:** Preferred dose and agent selection Table legend: $ Surgeons include trauma surgeons and general surgeons * Other specialties include orthopedic surgeons and neurosurgeons Abbreviations: Anti-Xa: serum anti-factor Xa concentrations, IVC: inferior vena cava, LMWH: low molecular weight heparin, PVTE-Px: pharmacological venous thromboembolism prophylaxis, RCT: randomized controlled trials, UFH: unfractionated heparin

|  | Total cohort<br>N/# of<br>responses (%) | Intensivists<br>N/# of<br>responses<br>(%) | Surgeons<sup>§</sup><br>N/# of<br>responses (%) | Other<br>specialties*<br>N/# of responses<br>(%) | P value |
| --- | --- | --- | --- | --- | --- |
| **Modality** |  |  |  |  |  |
| Mechanical if no contraindication (e.g. lower extremity injuries) | 4/63 (6.35) | 3/39 (7.69) | 0/15 (0) | 1/9 (11.11) | 0.4580 |
| Pharmacological | 22/63 (34.92) | 16/39 (41.03) | 4/15 (26.67) | 2/9 (22.22) |  |
| Combination of mechanical and pharmacological | 36/63 (57.14) | 19/39 (48.72) | 11/15 (73.33) | 6/9 (66.67) |  |
| IVC filter | 1/63 (1.59) | 1/39 (2.56) | 0/15 (0) | 0/9 (0) |  |
| **Pharmacological agents (select all that apply )** |  |  |  |  |  |
| Unfractionated heparin (UFH) | 24/102 (23.53) | 19/65 (29.23) | 5/23 (21.74) | 0/14 (0) | 0.0131 |
| Low molecular weight heparin (LMWH) | 53/ 102 (51.96) | 29/65 (44.62) | 15/23 (65.22) | 9/14 (64.29) | 0.1407 |
| Fondaparinux | 4/102 (3.92) | 4/ 65 (6.15) | 0 / 23 (0) | 0/ 14 (0) | 0.1575 |
| Warfarin | 1/102 (0.98) | 1/65 (1.54) | 0/23 (0) | 0/ 14 (0) | 0.6355 |
| **UFH dose** |  |  |  |  |  |
| 5000 units every 12 hours | 35/53 (66.03) | 24/34 (70.59) | 9/13 (69.23) | 2/6 (33.33) | 0.3151 |
| 5000 units every 8 hours | 14/53 (26.42) | 8/34 (23.53) | 4/13 (30.77) | 2/6 (33.33) |  |
| 2500 units every 12 hours | 1/53 (1.88) | 0/34 (0) | 0/13 (0) | 1/6 (16.67) |  |
| 2500 units every 8 hours | 1/53 (1.88) | 1/34 (2.94) | 0/13 (0) | 0/6 (0) |  |
| Undecided | 2/53 (3.77) | 1/34 (2.94) | 0/13 (0) | 1/6 (16.67) |  |
| **LMWH Dose** |  |  |  |  |  |
| 40 mg every 12 hours | 10/63 (15.87) | 5/39 (12.82) | 4/15 (26.67) | 1/9 (11.11) | 0.6509 |
| 30 mg every 12 hours | 5/63 (7.94) | 3/39 (7.69) | 1/15 (6.67) | 1/9 (11.11) |  |
| 40 mg once daily | 28/63 (44.44) | 19/39 (48.72) | 7/15 (46.67) | 2/9 (22.22) |  |
| Weight based dosing after discussion with clinical pharmacist | 19/63 (30.15) | 11/39 (28.21) | 3/15 (20) | 5/9 (55.56) |  |
| Undecided | 1/63 (1.59) | 1/39 (2.56) | 0/15 (0) | 0/9 (0) |  |
| **Utilizing anti-Xa level for adjustment of LMWH for PVTE-Px in trauma** |  |  |  |  |  |
| Yes | 4/62 (6.45) | 4/38 (10.53) | 0/15 (0) | 0/9 (0) | 0.1294 |
| **Interested in a multicenter prospective RCT** |  |  |  |  |  |
| Yes | 49/ 62 (79.03) | 31/38 (81.58) | 13/15 (86.67) | 5/9 (55.56) | 0.1993 |

Physicians perceptions to start early PVTE-Px per clinical scenarios varied **[Table 4: clinical scenarios**, **Table 5: clinical scenarios response categorized based on the specialties (intensivist vs. surgeons)]**. For TBI, 40.3% of respondents initiated PVTE-Px early within 24 hours, while 46.8 % were late initiators. When categorized based on their specialties (intensivists vs. surgeons) a significant proportion of surgeons would initiate PVTE-Px early in TBI within 24 hours (71.4 % and 37.5 %, respectively, P = 0.0271). For TBI with ICP monitor, 45.6 % of respondents will initiate PVTE-Px early within 24 hours compared to 29.8 % of late initiators if a repeated CT head was stable and showed no sign of progression; 83.3 % of them were surgeons and 52.6% were intensivists (P = 0.0479). Looking at the SCI scenario, 36.8% of respondents were early initiators without a repeated CT spine. Most surgeons would initiate PVTE-Px early within 24 hours but this was statistical insignificant (73.3 % and 50 %, respectively, P = 0.1136).

**Table 4:** Clinical scenarios Table legend: $ Surgeons include trauma surgeons, general surgeons * Other specialties include orthopedic surgeons and neurosurgeons # Early initiator defined as ≤24 hour and late initiator defined as >24 hour in clinical scenarios related to neurotrauma (TBI, TBI with ICP monitor, and SCI) ## Early initiator defined as ≤48 hour and late initiator defined as >48 hour in clinical scenarios related to NOR solid organ injuries (GSW through the Liver, grade IV splenic injury) † Wait until the ICP is within normal limits and wait until the ventriculostomy is removed Abbreviations: CT: computed tomography, NOR: non-operative, SCI: spinal cord injury, TBI: traumatic brain injury, GSW: gunshot wound, ICP: intracranial pressure

|  | Total cohort<br>N/# of<br>responses (%) | Intensivists<br>N/# of<br>responses (%) | Surgeons<sup>§</sup><br>N/# of responses<br>(%) | Other specialties *<br>N/# of responses<br>(%) | P value |
| --- | --- | --- | --- | --- | --- |
| **TBI Case<sup>#</sup>** |  |  |  |  |  |
| Early initiator without a repeated CT head | 4/62 (6.45) | 1/40 (2.50) | 3/14 (21.43) | 0/8 (0) | 0.1181 |
| Early initiator if a repeated CT head is stable | 25/62 (40.32) | 14/40 (35) | 7/14 (50) | 4/8 (50) |  |
| Late initiator without a repeated CT head | 4/62 (6.45) | 3/40 (7.50) | 1/14 (7.14) | 0/8 (0) |  |
| Late initiator if a repeated CT head is stable | 29/62 (46.77) | 22/40 (55.0) | 3/14 (21.43) | 4/8 (50) |  |
| **TBI Case with ICP monitor<sup>#</sup>** |  |  |  |  |  |
| Early initiator without a repeated CT head | 7/57 (12.28) | 6/38 (15.79) | 1/12 (8.33) | 0/7 (0) | 0.0800 |
| Early initiator if a repeated CT head is stable | 26/57 (45.61) | 14/38 (36.84) | 9/12 (75) | 3/7 (42.86) |  |
| Late initiator without a repeated CT head | 5/57 (8.77) | 5/38 (13.16) | 0/12 (0) | 0/7 (0) |  |
| Late initiator if a repeated CT head is stable | 17/57 (29.82) | 12/38 (31.58) | 1/12 (8.33) | 4/7 (57.14) |  |
| Other<sup>†</sup> | 2/57 (3.51) | 1/38 (2.63) | 1/12 (8.33) | 0/7 (0) |  |
| **SCI case<sup>#</sup>** |  |  |  |  |  |
| Early initiator without a repeated CT spine | 21/57 (36.84) | 13/37 (35.14) | 6/12 (50) | 2/8 (25) | 0.2166 |
| Early initiator if a repeated CT spine is stable | 15/57 (26.32) | 7/37 (18.92) | 5/12 (41.67) | 3/8 (37.50) |  |
| Late initiator without a repeated CT spine | 5/57 (8.77) | 4/37 (10.81) | 0/12 (0) | 1/8 (12.50) |  |
| Late initiator if a repeated CT spine is stable | 16/57 (28.07) | 13/37 (35.14) | 1/12 (8.33) | 2/8 (25) |  |
| **NOR solid organ injuries scenario 1: GSW through the Liver<sup>##</sup>** |  |  |  |  |  |
| Early initiator without a repeated CT abdomen | 28/57 (49.12) | 15/36 (41.67) | 12/14 (85.71) | 1/7 (14.29) | 0.0136 |
| Early initiator if a repeated CT abdomen is stable | 17/57 (29.82) | 12/36 (33.33) | 1/14 (7.14) | 4/7 (57.14) |  |
| Late initiator without a repeated CT abdomen | 4/57 (7.02) | 4/36 (11.11) | 0/14 (0) | 0/7 (0) |  |
| Late initiator if a repeated CT abdomen is stable | 8/57 (14.04) | 5/36 (13.89) | 1/14 (7.14) | 2/7 (28.57) |  |
| **NOR solid organ injuries scenario 2: grade IV splenic injury<sup>##</sup>** |  |  |  |  |  |
| Early initiator without a repeated CT abdomen | 20/55 (36.36) | 11/35 (31.43) | 8/13 (61.54) | 1/7 (14.29) | 0.1501 |
| Early initiator if a repeated CT abdomen is stable | 18/55 (32.73) | 11/35 (31.43) | 2/13 (15.38) | 5/7 (71.43) |  |
| Late initiator without a repeated CT abdomen | 5/55 (9.09) | 4/35 (11.43) | 1/13 (7.69) | 0/7 (0) |  |
| Late initiator if a repeated CT abdomen is stable | 12/55 (21.82) | 9/35 (25.71) | 2/13 (15.38) | 1/7 (14.29) |  |

**Table 5:** Clinical scenarios response categorized based on the specialties (intensivist vs. surgeons) Abbreviations: TBI: traumatic brain injury, SCI: spinal cord injury, GSW: gunshot wound, ICP: intracranial pressure

|  | Total | Early N (%) | Late N (%) | P value |
| --- | --- | --- | --- | --- |
| TBI |  |  |  |  |
| Intensivists | 40 | 15 (37.50) | 25 (62.50) | 0.0271 |
| Surgeons | 14 | 10 (71.43) | 4 (28.57) |  |
| TBI with ICP monitor |  |  |  |  |
| Intensivists | 38 | 20 (52.63) | 18 (47.37) | 0.0479 |
| Surgeons | 12 | 10 (83.33) | 2 (16.67) |  |
| SCI |  |  |  |  |
| Intensivists | 40 | 20 (50) | 20 (50) | 0.1136 |
| Surgeons | 15 | 11 (73.33) | 4 (26.67) |  |
| GSW to liver |  |  |  |  |
| Intensivists | 40 | 27 (67.50) | 13 (32.50) | 0.1355 |
| Surgeons | 15 | 13 (86.67) | 2 (13.33) |  |
| Grade IV splenic injury |  |  |  |  |
| Intensivists | 39 | 22 (56.41) | 17 (43.59) | 0.4888 |
| Surgeons | 15 | 10 (66.67) | 5 (33.33) |  |

With regards to NOR solid organ injuries [gunshot wound (GSW) to the liver and grade IV splenic injuries], 49.1% and 36.4% of the respondents respectively were considered early initiators within 48 hours without a repeat CT abdomen. The same trend was observed when they were categorized based on their specialties; higher number of surgeons started PVTE-Px early in NOR solid organ injuries compared to intensivists (P = 0.1355 and P = 0.4888 for GSW to the liver and grade IV splenic injury, respectively).

To better understand the predictors for the early initiation of PVTE-Px, we evaluated the perceived barriers as well as controlled for the injury trauma types among the respondents for each clinical scenario **[Table 6: Logistic regression models adjusted for the injury type and the perceived barriers, and supplementary table 3]**. The significant injury predictors for late initiation of PVTE-Px in TBI were MVA (OR 681, 95% CI: 1.7-274635, p=0.03), blunt injury (OR 640, 95% CI: 6.77-60627, p=0.005), and motorcycle (OR 41, 95%CI: 1-1729, p=0.05). While significant injury predictors for early initiation of PVTE-Px in TBI were pelvic fracture (OR 0.004, 95% CI: 8.6 e-5-0.23, p=0.007) and pedestrian (OR 0.01, 95% CI: 0.0001-0.717, p=0.0341). Two significant perceived barriers found to be predictors for late initiation of PVTE-Px in TBI were multiple surgical interventions (craniotomy, and ICP monitors insertions) (OR 31.9, 95% CI: 3.13-325, p=0.0035) and others barriers listed by respondents as free text [high International Normalized Ratio (INR), medico-legal issues, active bleeding or bleeding tendency, protocol-based practice, extent of trauma and presence of polytrauma, and presence of disseminated intravascular coagulation (DIC)] (OR 222, 95% CI: 1.36-36285, p=0.037). The most significant perceived barrier for early initiation of PVTE-Px in TBI with ICP monitor was severe head injuries (OR 0.33, 95% CI: 0.11-0.98, p=0.04).

**Table 6:**
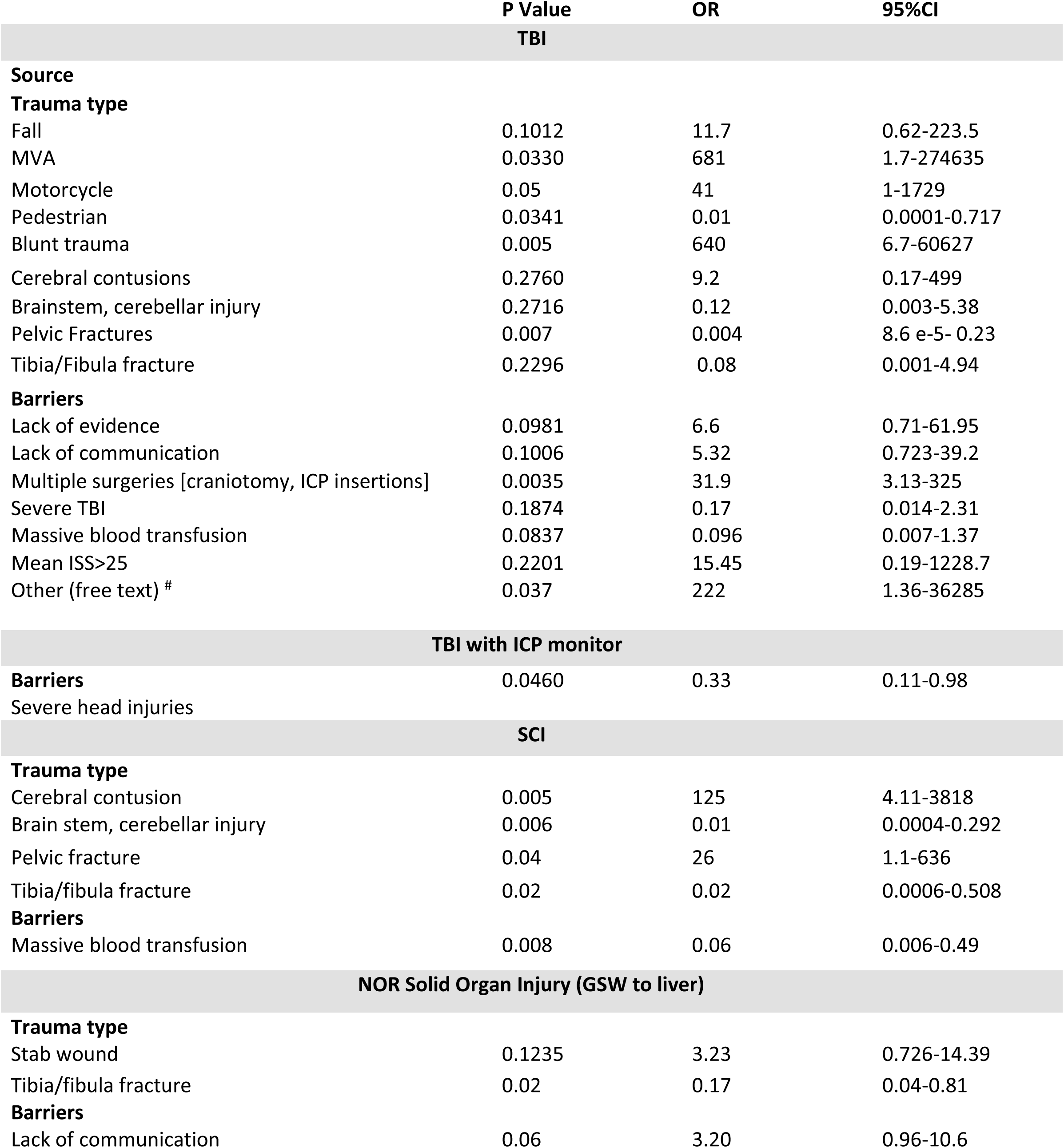
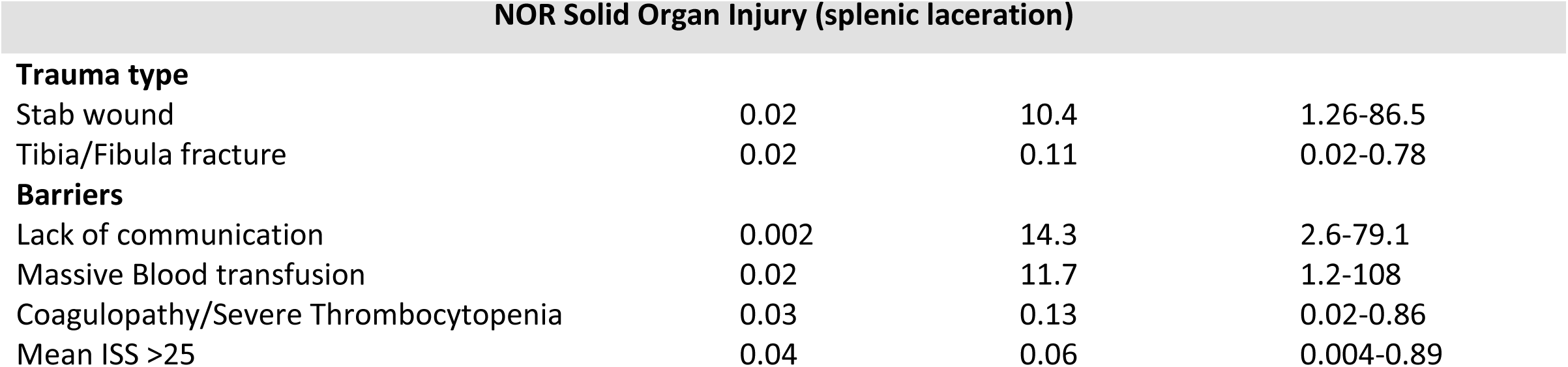
Logistic regression models adjusted for the injury type and the perceived barriers Table legend: The odds ratio (OR) is adjusted for injury type and perceived barriers. For P value < 0.05 and OR < 1 is a significant injury type predictor for early initiation or perceived barrier for early initiation, P value <0.05 and OR > 1 is a significant injury type predictor for late initiation or perceived barrier for late initiation. # Other (free text): high International Normalized Ratio (INR), medico-legal issues, active bleeding or bleeding tendency, protocol-based practice, extent of trauma and presence of polytrauma, and presence of disseminated intravascular coagulation (DIC). Abbreviation: OR: odds ratio, CI: confidence interval, TBI: traumatic brain injury, SCI: spinal cord injury, NOR : non-operative, GSW: gunshot wound, ICP: intracranial pressure, MVA: motor vehicle accident, ISS: injury severity score

|  | P Value | OR | 95%CI |
| --- | --- | --- | --- |
| **TBI** |  |  |  |
| **Source** |  |  |  |
| **Trauma type** |  |  |  |
| Fall | 0.1012 | 11.7 | 0.62-223.5 |
| MVA | 0.0330 | 681 | 1.7-274635 |
| Motorcycle | 0.05 | 41 | 1-1729 |
| Pedestrian | 0.0341 | 0.01 | 0.0001-0.717 |
| Blunt trauma | 0.005 | 640 | 6.7-60627 |
| Cerebral contusions | 0.2760 | 9.2 | 0.17-499 |
| Brainstem, cerebellar injury | 0.2716 | 0.12 | 0.003-5.38 |
| Pelvic Fractures | 0.007 | 0.004 | 8.6 e-5- 0.23 |
| Tibia/Fibula fracture | 0.2296 | 0.08 | 0.001-4.94 |
| **Barriers** |  |  |  |
| Lack of evidence | 0.0981 | 6.6 | 0.71-61.95 |
| Lack of communication | 0.1006 | 5.32 | 0.723-39.2 |
| Multiple surgeries [craniotomy, ICP insertions] | 0.0035 | 31.9 | 3.13-325 |
| Severe TBI | 0.1874 | 0.17 | 0.014-2.31 |
| Massive blood transfusion | 0.0837 | 0.096 | 0.007-1.37 |
| Mean ISS>25 | 0.2201 | 15.45 | 0.19-1228.7 |
| Other (free text) # | 0.037 | 222 | 1.36-36285 |
| **TBI with ICP monitor** |  |  |  |
| **Barriers** | 0.0460 | 0.33 | 0.11-0.98 |
| Severe head injuries |  |  |  |
| **SCI** |  |  |  |
| **Trauma type** |  |  |  |
| Cerebral contusion | 0.005 | 125 | 4.11-3818 |
| Brain stem, cerebellar injury | 0.006 | 0.01 | 0.0004-0.292 |
| Pelvic fracture | 0.04 | 26 | 1.1-636 |
| Tibia/fibula fracture | 0.02 | 0.02 | 0.0006-0.508 |
| **Barriers** |  |  |  |
| Massive blood transfusion | 0.008 | 0.06 | 0.006-0.49 |
| **NOR Solid Organ Injury (GSW to liver)** |  |  |  |
| **Trauma type** |  |  |  |
| Stab wound | 0.1235 | 3.23 | 0.726-14.39 |
| Tibia/fibula fracture | 0.02 | 0.17 | 0.04-0.81 |
| **Barriers** |  |  |  |
| Lack of communication | 0.06 | 3.20 | 0.96-10.6 |
| Trauma type |  |  |  |
| Stab wound | 0.02 | 10.4 | 1.26-86.5 |
| Tibia/Fibula fracture | 0.02 | 0.11 | 0.02-0.78 |
| Barriers |  |  |  |
| Lack of communication | 0.002 | 14.3 | 2.6-79.1 |
| Massive Blood transfusion | 0.02 | 11.7 | 1.2-108 |
| Coagulopathy/Severe Thrombocytopenia | 0.03 | 0.13 | 0.02-0.86 |
| Mean ISS >25 | 0.04 | 0.06 | 0.004-0.89 |

In case of SCI, the significant injury predictors for late initiation of PVTE-Px were cerebral contusion or laceration (OR 125, 95% CI: 4.11-3818, p=0.005) and pelvic fractures (OR 26, 95% CI: 1.1-636, p=0.04). While the significant injury predictors for early initiation of PVTE-Px were brainstem or cerebellar injuries (OR 0.01, 95%CI: 0.0004-0.292, p= 0.006), and tibia or fibula fractures (OR 0.02, 95% CI: 0.0006-0.508, p=0.02). One significant perceived barrier for early initiation of PVTE-Px was massive early transfusion and increasing blood transfusion requirements [>6 units of packed red blood cell (pRBC) within 12 hours of injury] (OR 0.06, 95% CI:0.006-0.49, p=0.008).

One significant injury predictor found to be associated with early initiation of PVTE-Px in GSW to the liver was tibia or fibula fracture (OR 0.17, 95% CI: 0.04-0.81. p=0.02). The injury predictors for late initiation of PVTE-Px in grade IV splenic injury were stab injuries (OR 10.4, 95% CI: 1.26-86.5, p=0.02) while the significant perceived barriers were lack of communication between services (surgical, critical care) (OR 14.3, 95% CI:2.6-79.1,p=0.002), massive early transfusion and increasing blood transfusion requirements (>6 units of pRBC within 12 hours of injury) (OR 11.7, 95% CI: 1.2-108, p=0.02). On the other hand, the injury predictor for early initiation of PVTE-Px in grade IV splenic injury was tibia or fibula fractures (OR 0.11, 95%CI: 0.02-0.78, p=0.02). Among the significant perceived barriers for early initiation, coagulopathy and severe thrombocytopenia (OR 0.13, 95% CI: 0.02-0.86, p=0.03), and the mean ISS >25 (OR 0.06, 95% CI: 0.004-0.89, p=0.04).

When unadjusted logistic regression was constructed for the perceived barriers, acute physiology and chronic health evaluation II (APACHE II) score > 25 were considered among the most significant perceived barrier for early initiation across all clinical scenarios [**Supplementary figure 1**].

A detailed response of clinical scenarios categorized based on the position, practicing years, and board certification years is available in supplementary material [**supplementary table 4,5,6**]. Notably, a higher percentage of consultants were classified as early initiators for the scenarios related to neurotrauma. For the scenario related to GSW to the liver, equal proportion of the consultants would start in 24 or 48 hours without a repeated CT abdomen. More conservative approach was noted in the scenario related to grade IV splenic injury. When responses were compared according to the number of practicing years, clinicians with ≤ 10 years of experience were classified as early initiators for the scenarios related to neurotrauma and NOR solid organ injuries.

## Discussion

It is known that the critically injured patient, while initially coagulopathic due to traumatic bleeding, often develop a hypercoagulable state, due to the systemic inflammatory response seen in post trauma patients, specifically an increase in C reactive proteins [37,38]. This phenomenon of a progression from coagulopathic to hypercoagulable state, coupled with the risks of ongoing hemorrhage in subgroups such as NOR managed solid organ injuries, or the TBI patients with ICH, make the PVTE-Px in trauma patients one of the most challenging issues. The study conducted by Nathens *et al* group describe the practice of PVTE-Px in the major trauma patient, 34/315 (11%) had a diagnosis of VTE, and one third of the events occurred in the first week after injury [39]. The proportion of patients developing VTEs increased significantly with delays in the initiation of PVTE-Px. Early prophylaxis was associated with a 5% risk of VTE, whereas delay beyond 4 days was associated with 3 times that risk (risk ratio, 3.0, 95% CI [1.4 – 6.5]). Therefore, it is evident that this delay is associated with a higher likelihood of VTE.

To the best of our knowledge, our study is the first study to evaluate and identify differences across specialties and practice patterns regarding the timing of PVTE-Px initiation in TBI, SCI, and NOR solid organ injuries. Variations in the reported clinical practice is not surprising given the inconsistent recommendations from guidelines regarding this topic. The need for future investigation is also highlighted by a recent literature.

In a retrospective study using the TQIP database that included over 3,000 patients with severe TBI, early initiation of PVTE-Px within 72 hours of injury halved the rate of VTE (PE odd ratio, 0.48 and DVT odds ratio, 0.51) compared with the rate in patients who received late prophylaxis (≥72 hours), without an increase in the risk of death or neurosurgical intervention for new or expanding ICH [40]. In another retrospective study in patients with ICH from TBI, early PVTE-Px did not alter the rate of VTE or ICH compared with patients treated after 48 hours [41]. Other studies suggest similar efficacy and safety when prophylaxis was started within 24 to 36 hours of blunt traumatic brain injury [42]. A randomized double-blind placebo-controlled study by Phelan *et al* evaluated the clinical difference between enoxaparin (30 mg twice daily) PVTE-Px started within 24-96 hours versus placebo in patients with TBI [43]. Sixty-two patients presenting with prespecified small TBI patterns and stable scans at 24 hours after injury were included. Findings showed that in-hospital VTE incidence rate was 0 % in enoxaparin arm vs 3.6 % in placebo arm. Radiographic TBI progression rates on the scans performed 48 hours after injury and 24 hours after start of treatment to be 5.9% (95% CI, 0.7-19.7%) for enoxaparin and 3.6% (95% CI, 0.1-18.3%) for placebo, a treatment effect difference of 2.3% (95% CI, −14.42-16.5%). The researchers concluded that the risk of TBI progression in patients with stable injuries at 24 hours who were initiated on enoxaparin PVTE-Px is minimal and were similar to those of placebo. Lately, a systematic review by Spano *et al* was conducted for studies evaluating the safety and efficacy of PVTE-Px in TBI patients [44]. Safety was defined as a stable hematoma or no increase in the size of intracranial bleed in patients who were initiated on PVTE-Px. Effectiveness was defined as a reduction in the overall incidence of VTE. Of the 21 studies included, 18 concluded that PVTE-Px in patients with follow-up head CT demonstrating a stable ICH does not lead to exacerbation of the bleed. Of the three studies that found PVTE-Px led to hemorrhage progression, one study included patients who required neurosurgical intervention, which may explain the high progression rate. About 14 studies demonstrated that prophylaxis between 24 and 72 hours is safe in patients with stable head CTs. In addition, four studies revealed that a timeframe for administration of PVTE-Px within 24 hours of injury was an acceptable time frame for stable patients. The definition for a stable CT scan was not given by the authors but presumably could be considered as CT imaging with an injury or bleed of the same size or less. The authors concluded that most studies demonstrated that early PVTE-Px administration is warranted for patients with stable repeat CT and associated with a decreased incidence of VTE in patients with TBI without an increase in ICH. Potential limitation of the studies included in this systematic review include single institution experience, some lacked control group, and high probability of type II error because of the limited sample size.

In our study, 40.3% of respondents initiated PVTE-Px early within 24 hours for TBI when a repeated CT head was stable and showed no sign of progression in TBI, while 46.8 % are late initiators. For TBI with ICP monitor, 45.6 % of respondents will initiate PVTE-Px early within 24 hours compared to 29.8 % late initiators if a repeated CT head was stable and showed no sign of progression. Moreover, a significant proportion of surgeons (trauma and general surgeons) would initiate PVTE-Px early in TBI and TBI with ICP monitor within 24 hours which is consistent with most former literature [53-55]. This finding is relatively in line with the ACS-TQIP recommendations for VTE prophylaxis in TBI patients [13]. These guidelines subcategorize patients into low, moderate, and high risk of hemorrhagic progression. Low-risk patients may be started on PVTE-Px if CT scans are stable at 24 hours. Moderate risk patients are classified as patients with a subdural or epidural hematoma of greater than 8 mm, contusion or intraventricular hemorrhage of greater than 2 cm, multiple contusions per lobe, or subarachnoid hemorrhage with an abnormal CT angiogram or patients whose imaging has evidence of progression at 24 hours and may be started on PVTE-Px with a stable head CT at 72 hours. Patients in the high risk category (i.e., hematoma progression at 72 hours, necessity of ICP monitoring, or craniotomy patients) should be considered for inferior vena cava filter placement to prevent VTE events.

In case of SCI, a single-center retrospective study of 1,432 patients with spinal fractures, 14% of whom had operative fixation, early PVTE-Px initiated within 24 hours was not associated with an increased risk of postoperative bleeding or epidural hematoma [45]. In our study, most respondents were early initiators within 24 hours, without a repeated CT spine, and a higher percentage of surgeons would initiate PVTE-Px early in SCI within 24 hours; however, this did not reach statistical significance compared to the intensivists. This is in line with 2020 WTA guideline and may be explained by the physicians’ perception of the high risk of SCI population for developing VTE because of increased venostasis, hypercoagulability, and frequently accompanying vascular injury [11,12,18].

Regarding NOR solid organ injuries, several retrospective studies and literature reviews of blunt solid-organ injuries demonstrated that early PVTE-Px initiation within 48 hours of admission reduces VTE without increasing bleeding [46,47,48]. A 4-year retrospective review from Eberle *et al* concluded that early initiation (<72 hours) of PVTE-Px in patients with blunt splenic, hepatic, and/or renal injuries was not associated with increased failure of nonoperative management or increased transfusion requirements [49]. Another 6-year retrospective study by Joseph *et al* including 116 patients with blunt abdominal solid organ injuries was conducted [50]. The authors used a propensity analysis for age, sex, blood pressure, Glasgow Coma Score (GCS), ISS, and type/grade of organ injury to compare early (<48 hours), intermediate (48–72 hours), and late (>72 hours) PVTE-Px. There was no significant correlation between early and post prophylaxis blood transfusion or need for operative intervention. Lastly, a 2-year (2013–2014) retrospective analysis of ACS-TQIP included all adult trauma patients (age ≥ 18 years) with blunt solid organ injuries who underwent NOR management of solid organ injuries [51]. A total of 36,187 patients were included and stratified into three groups based on timing of PVTE-Px (early, ≤48 hours of injury; late, >48 hours of injury; and no prophylaxis group). After controlling for confounders, patients receiving early PVTE-Px had lower rates of DVT (p = 0.01) and PE (p = 0.01) compared with the no prophylaxis and late PVTE-Px groups. There was no difference between the three groups regarding the post prophylaxis pRBC transfusions, failure of NOR management, and mortality. The results of this study suggested that in patients undergoing NOR management of blunt abdominal solid organ injuries, early initiation of PVTE-Px should be considered. However, it is noted that failure of NOR rate and post prophylaxis pRBC transfusion was significantly higher in the spleen and the kidney group relative to the liver group (p = 0.01) and higher grades of solid organ injuries (grade ≥ IV) were associated with higher failure of NOR rate in comparison to lower grades (p = 0.01). Our study results are consistent with this recent retrospective analysis where majority of the respondents were considered early initiators within 48 hours without a repeated CT abdomen in case of GSW to the liver. More disparate responses observed and seems more conservative in grade IV splenic injury.

In our study, most respondents used combined mechanical and PVTE-Px. Mechanical prophylaxis is used as an adjunct for high-risk patients (multiple, large, intracranial hematomas or contusions) if PVTE-Px is contraindicated but the presence of lower extremities fracture may limit their use. Unfortunately, the evidence supporting the efficacy of mechanical prophylaxis in this patient population is weak and compliance associated with these devices is relatively low [52,53]. This challenge is further magnified when one considers that many VTE events might occur within the first few days of injury, suggesting that if there is to be any benefit, prophylaxis should be initiated as early as possible. The Pneumatic Compression for Preventing Venous Thromboembolism (PREVENT) trial by Arabi and colleagues showed that among 2003 medical, surgical, and trauma intensive care units (ICU) patients, combined thromboprophylaxis was safe but did not result in a significantly lower incidence of proximal lower-limb DVT, PE, or other clinical outcomes than PVTE-Px alone [54]. These findings may highlight the importance of initiating the PVTE-Px as soon as the bleeding risk becomes acceptably low. However, It is worth mentioning that trauma patients in PREVENT trial represented only 8% of the trial population [55].

Notably, enoxaparin (LMWH) is selected by 52% of our respondents for critically ill trauma patients. The EAST guidelines recommend LMWH over UFH for high-risk patients—defined as having an ISS of 9 or greater [5]. The ACCP guidelines equivocally recommend UFH or LMWH over no prophylaxis but do not specify a preferential therapeutic agent [6]. Despite mixed results from studies comparing the efficacy of UFH vs. LMWH in trauma patients, most of our cohort and trauma centers preferentially select LMWH particularly in high-risk patients.

The preferential use of LMWH is supported by a recently published retrospective, propensity-matched cohort study comparing the incidence of PE and DVT between LMWH (n=37,960) and UFH (n=37,960) using data from the ACS-TQIP. The study suggested that LMWH use is associated with a 42% decreased risk of PE compared to UFH (1.4% vs. 2.4%) and a 28% decreased risk of DVT (3.9% vs. 5.4%) [3]. Additionally, serum anti-Xa concentrations have been proposed to monitor and adjust enoxaparin prophylactic dose because of the high risk of thrombosis and bleeding in trauma patients. End points include a peak serum concentration of 0.2–0.5 IU/mL and/or a trough serum concentration of 0.1– 0.2 IU/mL [56,57]. Some studies demonstrate a reduction in VTE with anti-Xa monitoring; others demonstrate no difference. In our study, small percentage of the respondent indicated the use of anti-Xa level to guide enoxaparin prophylactic dose. While dose adjusting to achieve a target anti-Xa may be an option, its utility is limited as a therapeutic range has not been clearly defined and an association between anti-Xa and VTE incidence or bleeding events has not been demonstrated yet [58, 59].

Moreover, a literature review suggested a weight-based dose of 0.5 mg/kg every 12 hours resulted in achieving a prophylactic anti-Xa concentration, but adequately powered studies are necessary to confirm the impact on VTE rate versus bleeding incidence [60,61]. In our study, 19/63 (30.15%) indicated weight-based dosing after discussion with clinical pharmacist. The ICU or ED clinical pharmacists are an increasingly important component of the trauma team, as evidenced by growth over the last decade. Study by Porter *et al* showed that pharmacist involvement with trauma resuscitation increased significantly in 71% of trauma centers practicing within the ED. Common bedside responsibilities include calculating dosages (96%), preparing medications (89%), providing medication information (79%), trauma team education (45%), research, ACS accreditation preparation, and others [62].

Looking to the study by Nathens *et al*, patients with severe head injury, those that received >6 units of pRBC within 12 hours of injury, and those without significant comorbidity are twice as likely to have PVTE-Px delayed beyond day 4. By contrast, the presence of a severe lower extremity fracture was associated with early PVTE-Px. Importantly, the presence of lower extremity fractures, a strong risk factor for VTE, did not modify the timing of prophylaxis among patients with severe head injuries, suggesting that the concern over bleeding took precedence [37]. It is worth mentioning that this study was limited to age ≥16 years, blunt trauma, arrival to hospital within 6 hours of injury, body region exclusive of the brain with an Abbreviated Injury Scale score (AIS) ≥2, and an intact cervical spinal cord while it excluded those with isolated severe head injuries or spinal cord injuries. Compared to Nathens *et al’s* results, we found that the significant injury predictors for early initiation of PVTE-Px in TBI were pelvic fracture while most significant perceived barrier for early initiation of PVTE-Px in TBI with ICP monitor was severe head injuries. In SCI, significant injury predictors for early initiation of PVTE-Px were brainstem or cerebellar injuries (OR 0.01, 95%CI: 0.0004-0.292, p= 0.006), and tibia or fibula fractures (OR 0.02, 95% CI: 0.0006-0.508, p=0.02). Tibia and fibula fracture were found to be among the significant injury predictor for early initiation in NOR solid organ injuries as well.

We also noticed that our study clinicians with < 10 years of experience were more likely to initiate PVTE-Px in line with the recently published literature (TBI with or without ICP monitors within 24 hours if a repeated CT head is stable, SCI within 24 hours without a repeated CT spine, and NOR solid organ injuries within 48 hours without a repeated CT abdomen) compared to those who practiced for > 10 years. We are unable to find the exact reasons for this but one speculation is that the juniors were more familiar with recent evidence based data having more recently completed their fellowship or residency training.

The fact that duplicate responses to the survey from the same participants were less likely in our study is considered one of our study strength. We asked in our initial soliciting message and subsequent reminders that all respondents be refrained from taking a duplicate survey. Another strengths of this study are a multicenter nature and inclusion of different specialties (intensivists, trauma surgeons, general surgeons, spine orthopedics, and neurosurgeons). Moreover, majority of our respondents were consultants with decent years of clinical experience. Furthermore, Saudi specific’ trauma research is important to develop, since much of the current trauma literature is derived from American and Canadian trauma centers that serve very different populations with different injury patterns. In addition, the Saudi government has launched the Saudi Vision 2030, analogous to the Healthy People 2020 program in the US (The Department of Health and Human Services 2010) [63]. One of the major goals of the Saudi Vision 2030 has been to promote preventive care to reduce disease, disability and injury (Council of Economic and Development Affairs 2016) [64]. Faced with the considerations presented, this study help us to improve our understanding and assess the practice patterns regarding the dose and timing of PVTE-Px initiation in TBI, SCI, and NOR solid organ injuries. The findings from this study will provide a basis for a future research that will assess this important preventive measure on clinically relevant outcomes and this will ultimately improve population health in our country.

However, the nature of the survey-based investigation inherently carries limitations which warrant discussion. The small sample size reflects the relatively small number of Trauma Centers located across Saudi Arabia, as compared to the wide distribution across the United States and Canada. Therefore, our study is possibly underpowered to detect a difference and statistical difference might be due to chance. Moreover, this was a cross-sectional study limiting the ability to examine causal relationships between specialties perceptions and practice patterns regarding timing of initiating PVTE-Px in TBI, SCI and NOR solid organ injuries patients. Although great care was taken to design questions for the survey, the clinical scenarios might not be detailed enough to guide the respondents and the available answer choices were not completely reflective of their preferred management. Therefore, it is quite possible that some of the variations seen in the responses to this survey may be due to difficulty in understanding the questions or unavailability of management of choice by the respondents, but not because of the variation in practice. Although we made every effort based on established principles to increase the response rate, our response rate was only 29 % which is lower than the proposed range of 50% and this may limit the generalizability of the conclusions. Yet, recent evidence indicates that a higher survey response rate does not necessarily improve the accuracy compared with surveys with lower response rates [65,66]. Overall, most respondents in our study worked in central region of Saudi Arabia and the conclusions may not be generalizable to other centers. It would be interesting to examine variation in early PVTE-Px decisions in other regions. Unfortunately, the few number of physicians in other regions did not allow for subgroup analyses. Moreover, memory could be a poor data capture method, and could lead to erroneous, misleading results. Finally, clinical studies of prophylactic measures in trauma patients with multiple injuries (poly-trauma) are limited thus studies on this heterogeneous population requires a very large study size.

Overall, we believe it is relevant to share our experience to set the stage for a for a larger multicenter, randomized controlled study powered enough to answer many questions regarding early PVTE-Px to refine existing care guidelines, better direct the physician at the bedside, and assess the efficacy and safety of different dosing accounting for ARC phenomenon. Further research is necessary to better define the scope and potential benefits of ICU or ED clinical pharmacist involvement in trauma resuscitation and hospital trauma programs. However, the logical next step would be the proposal of a national evidence-based VTE prophylaxis guidelines in our trauma population and the development of a national trauma registry [67]. While awaiting more robust data from multicenter randomized controlled trials, clinicians should exercise their judgment on a case-by-case basis regarding the timing of PVTE-Px in trauma patients and balancing the risks of worsening hemorrhage and development of VTE considering other factors such as the severity of injury, associated injuries, timing of surgical intervention, as well as other predisposing risk factors for VTE.

## Conclusion

Variations were observed in PVTE-Px initiation time influenced by trauma type. Nonetheless, our study results are relatively in line with the recent evidence-based clinical literature regarding the timing of PVTE-Px in TBI and NOR solid organ injuries. Our findings suggested half of the respondents used combined mechanical and PVTE-Px, and preferred LMWH in a standard prophylactic dose possibly indicating limited awareness of ARC and utilization of anti-Xa level. More robust data from randomized controlled trials are needed to establish the safety and efficacy of early PVTE-Px in adult trauma patients with different dosing regimens accounting for ARC phenomenon. The use of clinicians’ judgment is recommended on a case-by-case basis regarding the timing and dose of PVTE-Px in trauma patients and balancing the risks of worsening hemorrhage and development of VTE.

## Data Availability

available with corresponding author upon request

## List of Abbreviation

(AIS): Abbreviated Injury Scale1
(APACHE II): Acute Physiology and Chronic Health Evaluation II
(ACCP): American College of Chest Physicians
(ACS): American College of Surgeons
(TQIP): Trauma Quality Improvement Program
(AAST): American Association for the Surgery of Trauma
(ARC): Augmented renal clearance
(CRC): Clinical Research Committee
(CI): Confidence Interval
(DVT): Deep venous thrombosis
(ED): Emergency department
(EAST): Eastern Association for the Surgery of Trauma
(RAP): Greenfield Risk Assessment Profile
(GSW): Gunshot Wound
(GCS): Glasgow Coma Score
(ICH): Intracranial Hemorrhage
(ICU): Intensive care unit
(ICP): Intracranial pressure
(ISS): Injury Severity Score
(INR): International Normalized Ratio
(IRB): Institutional review board
(LMWH): Low molecular weight heparin
(NOR) solid organ injuries: Non-operative
(OR): Odds Ratio
(PVTE-Px): Pharmacological venous thromboembolism prophylaxis
(PRBCs): Packed red blood cells
(REC): Research Ethics Committee
(RAC): Research Advisory Council
(REDCap): Research Electronic Data Capture
(anti-Xa): Serum anti-factor-Xa concentrations
(SCI): Spinal cord injury
(TBI): Traumatic brain injury
(UFH): Unfractionated heparin
(VTE): Venous thromboembolism
(WTA): Western Trauma Association

## Acknowledgment

We thank Dr. Eman Abdulkareem Bakhsh, Consultant Neuroradiology at King Fahad Medical City Riyadh, Mohammed Abdulkareem Bakhsh, Orthopedic surgeon at King Fahad General Hospital Jeddah, Dr. Abdulwahed Barnawi, Neurosurgeon / spinal surgeon at PSMMC Riyadh, Sumayah Abunayyan Administrative Assistant at Saudi Association of Neurological Surgery (SANS), and Khadija Magadi; KFSHRC health outreach coordinator for their support and help in survey distribution. We sincerely thank all the survey respondents and medical staff at KFSHRC who participated in the survey and their support for this study particularly (Ashraf Tarifi, Eiad Kseibi, Mohammed Jamil, Khurshid Sayed, Omar Al Nafel, Kareema, Mohammed Jamil, Mohsen Khalil, Ali Zeeshan, Aaqib Jahangir, Abujazar Mohammed, Ahmed Fouad, Abid Butt, Hamad Alansari, Ali Aljanoubi, Ali Zeeshan, and Nabil Abouchala). We are also thankful for Saudi Critical Care Society for providing feedback of the study proposal, and Areej Alfattani for providing help in biostatistical part of manuscript.

## Tables and Table legends

## Supplementary materials

**supplementary table 1:** Summary of clinical practice guideline recommendations

Abbreviation: CT: computed tomography, EAST : Eastern Association for the Surgery of Trauma, ACS: American College of Surgeons TQIP: Trauma Quality Improvement Program, TBI: traumatic brain injury, SCI: spinal cord injury, NOR : non-operative, LMWH : low-molecular-weight heparin, UFH : unfractionated heparin, ICH: Intracranial hemorrhage, WTA: Western Trauma Association.

**supplementary table 2:** Summary of Enoxaparin thromboprophylaxis dosing in trauma patients Abbreviations: ICU: intensive care unit, LMWH : low-molecular-weight heparin, Anti-Xa: serum anti-factor Xa concentrations, DVT: deep vein thrombosis, VTE: venous thromboembolism

**Supplementary file 1**: Survey questions

**supplementary table 3:** Perceived barriers for early initiation of PVTE-Px

† Respondents could select multiple answers (select all that apply question)

$ Surgeons includes trauma surgeons, general surgeons

* Other specialties includes orthopedic surgeons, neurosurgeons

\# Other (free text): High INR, medicolegal issues, active bleeding or bleeding tendency, protocol-based practice, extent of trauma and presence of polytrauma, presence of disseminated intravascular coagulation (DIC)

Abbreviations: APACHE II: Acute Physiology and Chronic Health Evaluation II, ICP: intracranial pressure, PVTE-Px: pharmacological venous thromboembolism prophylaxis, pRBC: packed red blood cells

**Supplementary Figure 1:** Forest plots for unadjusted perceived barriers for early initiation of PVTE-Px Figure legend: The odds ratio (OR) is unadjusted and represents the odds of perceived barriers only. Abbreviations: OR: odds ratio, TBI: traumatic brain injury, ICP: intracranial pressure, SCI: spinal cord injury, SOI: solid organ injury, GSW: gunshot wound, APACHE II: Acute Physiology and Chronic Health Evaluation II, PVTE-Px: pharmacological venous thromboembolism prophylaxis

**supplementary table 4:** comparison between different responses in clinical scenarios according to the positions

Abbreviations: CT: computed tomography, TBI: traumatic brain injury, SCI: spinal cord injury, NOR: non-operative, GSW: gunshot wound, ICP: intracranial pressure

**supplementary table 5**: comparison of different responses in clinical scenarios according to practicing years

Abbreviations: CT: computed tomography, TBI: traumatic brain injury, SCI: Spinal Cord Injury, NOR: non-operative, GSW: gunshot wound, ICP: intracranial pressure

**supplementary table 6** : comparison of different responses in clinical scenarios according to the board certification

Abbreviations: CT: computed tomography, TBI: traumatic brain injury, SCI: Spinal Cord Injury, NOR: non-operative, GSW: gunshot wound, ICP: intracranial pressure

## References

1 Shackford SR, Davis JW, Hollingsworth-Fridlung P, Brewer NS, Hoyt DB, Mackersie RC. Venous thromboembolism in patients with major trauma. Am J Surg. 1990;159:365–9.

2 Geerts WH, Code KI, Jay RM, Chen E, Szalai JP. A prospective study of venous thromboembolism after major trauma. N Engl J Med. 1994;331:1601–6.

3 Byrne JP, Geerts W, Mason SA, Gomez D, Hoeft C, Murphy R, et al. Effectiveness of low-molecular-weight heparin versus unfractionated heparin to prevent pulmonary embolism following major trauma: a propensity-matched analysis. J Trauma Acute Care Surg. 2017;82:252–62.

4 Gearhart MM, Luchette FA, Proctor MC, Lutomski DM, Witsken C, James L et al. The risk assessment profile score identifies trauma patients at risk for deep vein thrombosis. Surgery 2000;128(4):631–40.

5 Rogers FB, Cipolle MD, Velmahos G, Rozycki G, Luchette FA. Practice management guideline for the prevention of venous thromboembolism in trauma patients: the EAST practice management guidelines work group. J Trauma 2002;53:142–64.

6 Guyatt GH, Akl EA, Crowther M, Gutterman DD, Schuünemann HJ. American College of Chest Physicians Antithrombotic Therapy and Prevention of Thrombosis Panel Executive summary. Antithrombotic therapy and prevention of thrombosis, 9th ed: American College of Chest Physicians evidence-based clinical practice guidelines. Chest 2012; 141 :7S–42S.

7 Barrera LM, Pereal P, Ker K, Cirocchi R, Farinella E, Uribe CH. Thromboprophylaxis for trauma patients. Cochrane Database Sys Rev 2013; 3:CD008303.

8 Taylor CA, Bell JM, Breiding MJ, Xu. Traumatic brain injury-related ED visits, hospitalizations, and deaths: United States, 2007 and 2013. MMWR 2017;66:1–16.

9 Bawazeer M, Amer M, Maghrabi K, Amin R, Vol ED, Hijazi M. Timing and dose of pharmacological thromboprophylaxis in adult trauma patients. Saudi Crit Care J. 2020;4:12–4.

10 Carney N, Totten AM, O’Reilly C. Guidelines for the Management of Severe Traumatic Brain Injury, Fourth Edition. Brain Trauma foundation. Neurosurgery. 2017 ;80:6–15.

11 Nyquist P, Jichici D, Bautista C, Burns J, Chhangani S, DeFilippis M, et al. Prophylaxis of venous thrombosis in neurocritical care patients: an executive summary of evidence-based guidelines: a statement for healthcare professionals from the Neurocritical Care Society and Society of Critical Care Medicine. Crit Care Med 2017;42:476–9

12 Sauro KM, Soo A, Kramer A, Couillard P. Venous Thromboembolism Prophylaxis in Neurocritical Care Patients: Are Current Practices, Best Practices?. Neurocrit Care. 2019;30:355–63

13 ACS TQIP Best Practice Guidelines. Management of Traumatic Brain Injury. Available at https://www.facs.org/quality-programs/trauma/tqp/center-programs/tqip/best-practice. Accessed November 14th, 2019

14 Ley EJ, Brown CV, Moore EE, Sava JA, Peck KA, Ciesla DJ, et al. Updated guidelines to reduce venous thromboembolism in trauma patients: A Western Trauma Association critical decisions algorithm. J Trauma Acute Care Surg. 2020.

15 Strollo BP, Bennett GJ, Chopko MS, Guo WA. Timing of venous thromboembolism chemoprophylaxis after traumatic brain injury. J Crit Care. 2018;43:75–80.

16 Dengler BA, Mendez-Gomez P, Chavez A, Avila L, Michalek J, Hernandez B. Safety of chemical DVT prophylaxis in severe traumatic brain injury with invasive monitoring devices. Neurocrit Care 2016.

17 Yao JS. Deep vein thrombosis in spinal cord-injured patients. Evaluation and assessment. Chest. 1992;102:645S–8S.

18 Tracy BM, Dunne JR,Oneal CM, Clayton E. Venous thromboembolism prophylaxis in neurosurgical trauma patients. J Surg Res. 2016;205:221–7

19 Walters BC. Methodology of the guidelines for the management of acute cervical spine and spinal cord injuries. Neurosurgery 2013;72:17–21.

20 The American Association for the Surgery of Trauma. Injury Scoring Scale. Accessed Nov 11, 2019 http://www.aast.org/Library/TraumaTools/InjuryScoringScales.aspx

21 Schellenberg M, Benjamin E, Piccinini A, Inaba K, Demetriades D. Gunshot wounds to the liver: No longer a mandatory operation. J Trauma Acute Care Surg. 2019;87:350–5.

22 Schellenberg M, Benjamin E1, Piccinini A, Inaba K1, Demetriades D. Selective Nonoperative Management of Renal Gunshot Wounds. J Trauma Acute Care Surg. 2019 Aug 14.

23 Van PY, Schreiber MA. Contemporary thromboprophylaxis of trauma patients. Curr Opin Crit Care. 2016;22:607–12

24 Stassen NA, Bhullar I, Cheng JD, Crandall M, Friese R, Guillamondegui O, et al. Nonoperative management of blunt hepatic injury: an Eastern Association of the Surgery of Trauma practice management guideline. J Trauma 2012; 73:S288–S293

25 Stassen NA, Bhullar I, Cheng JD, Crandall ML, Friese RS, Guillamondegui OD, et al. Selective nonoperative management of blunt splenic injury: an Eastern Association of the Surgery of Trauma practice management guideline. J Trauma 2012; 73:S294–S300.

26 Jacobs BN, Cain-Nielsen AH, Jakubus JL, Mikhail JN, Fath JJ, Regenbogen SE, et al. Unfractionated heparin versus lowmolecular-weight heparin for venous thromboembolism prophylaxis in trauma. J Trauma Acute Care Surg. 2017;83:151.

27 Arnold JD, Dart BW, Barker DE, Maxwell RA, Burkholder HC, Mejia VA, et al. Unfractionated heparin three times a day versus enoxaparin in the prevention of deep vein thrombosis in trauma patients. Am Surg. 2010;76:563–70.

28 Cook AM, Hatton-Kolpek J. Augmented Renal Clearance. Pharmacotherapy 2019;39:346–54.

29 Malinoski D, Jafari F, Ewing T, Ardary C, Conniff H, Baje M, et al. Standard therapeutic enoxaparin dosing leads to inadequate anti-Xa levels and increased deep venous thrombosis rates in critically ill trauma and surgical patients. J Trauma. 2010;68:874–80.

30 Kopelman TR, O’Neil PJ, Pieri PG, Salomone JP, Hall ST, Quan A, et al. Alternative dosing of prophylactic enoxaparin in the trauma patient: is more the answer? Am J Surg. 2013;206:911–6.

31 Nunez JM, Becher RD, Rebo GJ, Farrah JP, Borgerding EM, Stirparo JJ, et al. Prospective evaluation of weight-based prophylactic enoxaparin dosing in critically ill trauma patients: adequacy of antiXa levels is improved. Am Surg. 2015;81:605–9.

32 Berndtson AE, Costantini TW, Lane J, Box K, Coimbra K. If some is good more is better: an enoxaparin dosing strategy to improve pharmacologic venous thromboembolism prophylaxis. J Trauma Acute Care Surg. 2016;81:1095–100.

33 Costantini TW, Min E, Box K, Tran V, Winfield RD, Fortlage D, et al. Dose adjusting enoxaparin is necessary to achieve adequate venous thromboembolism prophylaxis in trauma patients. J Trauma Acute Care Surg. 2013;74:128–35.

34 Rostas JW, Brevard SB, Ahmed N, Allen J, Thacker D, Replogle WH, et al. Standard dosing of enoxaparin for venous thromboembolism prophylaxis is not sufficient for most patients within a trauma intensive care unit. Am Surg. 2015;81:889–92.

35 Burns KE, Duffett M, Kho ME, Meade MO, Adhikari NK, Sinuff T, et al. A guide for the design and conduct of self-administered surveys of clinicians. CMAJ 2008;179: 245–52.

36 von Elm E, Altman DG, Egger M, Pocock SJ, Gøtzsche PC, Vandenbroucke JP, et al. The Strengthening the Reporting of Observational Studies in Epidemiology (STROBE) statement: guidelines for reporting observational studies. Int J Surg Lond Engl. 2014;12:1495–9.

37 Cohen MJ, West M. Acute traumatic coagulopathy: from endogenous acute coagulopathy to systemic acquired coagulopathy and back. J Trauma 2011;70:S47–49.

38 Differding JA, Underwood SJ, Van PY, Khaki RA, Spoerke NJ, Schreiber MA. Trauma induces a hypercoagulable state that is resistant to hypothermia as measured by thrombelastogram. Am J Surg 2011;201:587–91.

39 Nathens AB, McMurray MK, Cuschieri J, Durr EA, Moore EE, Bankey PE, et al. The practice of venous thromboembolism prophylaxis in the major trauma patient. J Trauma. 2007;62:557–62.

40 Byrne JP, Mason SA, Gomez D, Hoeft C, Subacius H, Xiong W, et al. Timing of pharmacologic venous thromboembolism prophylaxis in severe traumatic brain injury: a propensity-matched cohort study. J Am Coll Surg. 2016; 223:621.

41 Frisoli F, Huang PP, Frangos S. 180 Early Deep Vein Thrombosis Chemoprophylaxis in Traumatic Brain Injury. Neurosurgery 2016; 63 :171.

42 Norwood SH, Berne JD, Rowe SA, Villarreal DH, Ledlie JT. Early venous thromboembolism prophylaxis with enoxaparin in patients with blunt traumatic brain injury. J Trauma 2008;65:1021–7.

43 Phelan HA, Wolf SE, Norwood SH, Aldy K, Brakenridge SC, Eastman AL, et al. A randomized, double-blinded, placebo-controlled pilot trial of anticoagulation in low-risk traumatic brain injury: the Delayed Versus Early Enoxaparin Prophylaxis I (DEEP I) study. J Trauma Acute Care Surg. 2012;73:1434–41

44 Spano PJ 2nd, Shaikh S, Boneva D, Hai S, McKenney M, Elkbuli A. Anticoagulant chemoprophylaxis in patients with traumatic brain injuries: A systematic review. J Trauma Acute Care Surg. 2020;88:454–460.

45 Kim DY, Kobayashi L, Chang D, Fortlage D, Coimbra R. Early pharmacological venous thromboembolism prophylaxis is safe after operative fixation of traumatic spine fractures. Spine (Phila Pa 1976) 2015;40:299.

46 Kwok AM, Davis JW, Dirks RC, Wolfe MM, Kaups KL et al. Time is now: venous thromboembolism prophylaxis in blunt splenic injury. Am J Surg. 2016;212:1231–6.

47 Murphy PB, Sothilingam N, Stewart TC, Batey B, Moffat B, Gray DK, et al. Very early initiation of chemical venous thromboembolism prophylaxis after blunt solid organ injury is safe. Can J Surg. 2016;59:118–22.

48 Rostas JW, Manley J, Gonzalez RP, Brevard SB, Ahmed N, Frotan MA, et al. The safety of low molecular-weight heparin after blunt liver and spleen injuries. Am J Surg. 2015;210:31–4.

49 Eberle BM, Schnuriger B, Inaba K, Cestero R, Kobayashi L, Barmparas G, et al. Thromboembolic prophylaxis with low-molecular-weight heparin in patients with blunt solid abdominal organ injuries undergoing nonoperative management: current practice and outcomes. J Trauma 2011;70:141–7.

50 Joseph B, Pandit V, Harrison C, Lubin D, Kulvatunyou N, Zangbar B, et al. Early thromboembolic prophylaxis in patients with blunt solid abdominal organ injuries undergoing nonoperative management: is it safe? Am J Surg. 2015; 209:194–8.

51 Skarupa D, Hanna K, Zeeshan M, Madbak F, Hamidi M, Haddadin Z, et al. Is early chemical thromboprophylaxis in patients with solid organ injury a solid decision?. J Trauma Acute Care Surg. 2019;87:1104–12.

52 Gould MK, Garcia DA, Wren SM, Karanicolas PJ, Arcelus JI, Heit JA, et al. Prevention of VTE in nonorthopedic surgical patients: antithrombotic therapy and prevention of thrombosis, 9th ed: American College of Chest Physicians Evidence-Based Clinical Practice Guidelines. Chest 2012;141:e227S.

53 Haut ER, Garcia LJ, Shihab HM, Brotman DJ, Stevens KA, Sharma R, et al. The effectiveness of prophylactic inferior vena cava filters in trauma patients. JAMA Surg. 2014;149:194–202.

54 Arabi YM, Al-Hameed F, Burns KEA, Mehta S, Alsolamy SJ, Alshahrani MS, et al. Adjunctive intermittent pneumatic compression for venous thromboprophylaxis. N Engl J Med. 2019;380:1305–15.

55 Lauzier F, Douketis JD, Cook DJ. A Device on Trial -Intermittent Pneumatic Compression in Critical Care. N Engl J Med. 2019;380:1367–8.

56 Haas CJ, Helsen JL, Raghavendra K, Mihalko W, Beres J, Ma Q, et al. Pharmacokinetics and pharmacodynamics of enoxaparin in multiple trauma patients. J Trauma 2005;59:1336–43.

57 Malinoski D, Jafari F, Ewing T, Ardary C, Conniff H, Baje M, et al. Standard prophy¬lactic enoxaparin dosing leads to inadequate anti-Xa levels and increased deep venous thrombosis rates in critically ill trauma and surgical patients. J Trauma 2010;68:874–80.

58 Vandiver JW, Ritz LI, Lalama JT. Chemical prophylaxis to prevent venous thromboembolism in morbid obesity: literature review and dosing recommendations. J Thromb Thrombolysis 2016;41:475–81.

59 Shaikh S, Boneva D, Hai S, McKenney M, Elkbuli A. Venous thromboembolism chemoprophylaxis regimens in trauma and surgery patients with obesity: A systematic review. J Trauma Acute Care Surg. 2020 ;88:522–35.

60 Chapman SA, Irwin ED, Reicks P, Beilman GJ. Non-weight-based enoxaparin dosing subtherapeutic in trauma patients. J Surg Res 2016;201:181–7.

61 Kopelman TR, Walters JW, Bogert JN, Basharat U, Pieri PG, Davis KM, et al. Goal directed enoxaparin dosing provides superior chemoprophylaxis against vein thrombosis. Injury 2017;48:1088–92

62 Porter BA, Zaeem M, Hewes PD, Hale LS, Jones CMC, Gestring ML, et al. Pharmacist involvement in trauma resuscitation across the United States: A 10-year follow-up survey. Am J Health Syst Pharm. 2019 ;76:1226–30.

63 Department of Health and Human Services. Healthy people 2020. 2010. https://www.healthypeople.gov. Accessed May 10 2020.

64 National Transformation Program. Saudi Vision 2030. 2016. http://vision2030.gov.sa/sites/default/files/NTP_En.pdf Accessed May 10 2020.

65 Curtin R, Presser S, Singer E. The effects of response rate changes on the index of consumer sentiment. Public opinion quarterly. 2000;64:413–28.

66 Holbrook AL, Krosnick JA, Pfent A. The causes and consequences of response rates in surveys by the news media and government contractor survey research firms. Advances in telephone survey methodology. 2008;1:499–528.

67 Alghnam S, Alkelya M, Al-Bedah K, Al-Enazi S. Burden of traumatic injuries in Saudi Arabia: lessons from a major trauma registry in Riyadh, Saudi Arabia. Ann Saudi Med. 2014;34:291–6.

