## supplemental file for "Timing and Dose of Pharmacological Thromboprophylaxis in Adult Trauma Patients: Perceptions, Barriers, and Experience of Saudi Arabia Practicing Physicians"

**Online Supplementary materials, tables and figures**

**Supplementary Table 1: Summary of clinical practice guideline recommendations**

<table>
<tr><th colspan="4">TBI Guidelines</th></tr>
<tr><th>Brain Trauma Foundation [10]</th><th>2017 Neurocritical Care Society [11,12]</th><th>ACS-TQIP [13]</th><th>2020 WTA Guideline [14]</th></tr>
<tr><td>Considered if the brain injury is stable and the benefit outweighs the risk of ↑ ICH</td><td>LMWH or UFH within 24 hours of TBI and 24 hours post craniotomy</td><td>Modified Berne-Norwood criteria start 24 hours (if low risk) , 72 hours (if moderate risk) with stable head CT scan<br>High-risk patients, namely, those needing neurosurgical intervention or progression at 72 hours, should be considered for a retrievable inferior vena cava filter</td><td>24 hours after injury for patients with stable CT and nearly all should receive within 72 hours of the time of injury . No consensus regarding the safety in the presence of ICP monitors or post craniotomy</td></tr>
<tr><th colspan="4">SCI Guidelines</th></tr>
<tr><th>2017 neurocritical care society [11,12]</th><th colspan="2">American Association of Neurological Surgeons (AANS) and Congress of Neurological Surgeons (CNS) [18]</th><th>2020 WTA Guideline [14]</th></tr>
<tr><td colspan="3">Start LMWH or UFH within 72 hours of admission with SCI</td><td>As soon as possible after a spine surgery or any spinal injury. Regimens can be initiated preoperatively or immediately after operative fixation (within 48 hours of operative fixation of traumatic spine fractures)</td></tr>
<tr><th colspan="4">NOR Solid Organ Injury (liver, spleen, and kidney) Guidelines</th></tr>
<tr><th colspan="3">2002 EAST [5, 23,24,47]</th><th>2020 WTA Guideline [14]</th></tr>
<tr><td colspan="3">No literature consensus about safe initiation time. Eberle et al: early (<3days) did not ↑ failure rates or blood transfusion</td><td>within 12 to 24 hours appeared to be safe across moderate American Association for the Surgery of Trauma (AAST) injury grades. Caution with grades IV and V injuries</td></tr>
</table>

**Supplementary Table 2: Summary of Enoxaparin thromboprophylaxis dosing in trauma patients**

| Authors | Study type | Intervention | Patients | Outcomes |
|---|---|---|---|---|
| Kopelman et al [29] | retrospective chart review | enoxaparin 40 mg every 12 hours ↑ goal anti-Xa levels vs 30 mg every 12 hours. All patients utilized sequential compression devices (SCDs) unless contraindicated as a result of fractured extremities | 124 trauma patients who were admitted to an urban, level 1 trauma center | Higher dosing of enoxaparin was associated with improved anti-Xa levels, this did not cause a statistically significant decrease in VTE in this small trial. 1 patient experienced event of bleeding (type of bleeding not defined) |
| Nunez et al [30] | prospective, nonrandomized cohort study | weight-based LMWH group ( 0.6 mg/kg dose every 12 hours) vs historical control group (standard 30-mg every 12 hours dosing regimen) | 37 trauma ICU patients | Weight-based dosing group had significantly more anti-Xa troughs within goal after the third dose and within 7 days (P <0.0001). DVT occurred in 1 patient in the weight-based group who had sustained a penetrating trauma to that extremity. Bleeding occurred in 1 patient in the weight-based group at the site of a removed central line |
| Berndtson et al [31] | prospective cohort study | Pre-intervention group (historical patients received enoxaparin 30 mg every 12 hours) and was compared with new dosing protocol (50 to 60 kg received 30 mg every 12 hours, 61 to 99 kg received 40 mg every 12 hours, and ≥100 kg received 50 mg every 12 hours) | obese trauma patients. | New dosing regimen had a higher rate of anti-Xa peak levels, lower VTE incidence. Bleeding events were similar, with 1 stable retroperitoneal hematoma developed new onset hemorrhage requiring angioembolization and blood product transfusion. |
| Costantini et al [32] | prospective cohort study | All patients initially received enoxaparin 30 mg every 12 hours, with doses adjusted per protocol to reach the goal anti-Xa peak level (<0.2 IU/mL, increase dose by 10 mg). | 61 level 1 academic trauma center | 43 (70.5%) were found to have sub-therapeutic initial anti-Xa levels on the standard regimen of enoxaparin 30 mg every 12 hours. |
| Rostas et al [33] | prospective cohort study | received enoxaparin 30 mg every 12 hours, adjusted per protocol until the anti-Xa trough was at goal (<0.1 IU/ mL, increase each dose by 10 mg). | 34 trauma ICU patients | To reach the goal of anti-Xa level, the dose had to be increased for 16 of the 28 patients included in this analysis. Four patients required 40 mg every 12 hours, 3 required 50 mg every 12 hours, 5 required 60 mg every 12 hours, 3 required 70 mg every 12 hours, and 1 patient required 150 mg every 12 hours. Adverse events were not reported. |

**Supplementary file 1, Survey questions**

**Demographic Data: Practitioner and Practice Site information**

What is your specialty? (Select all that apply)

- Intensivist
- Trauma Surgeon
- General Surgeon
- Orthopedic Surgeon
- Neurosurgeon

What is your position

- Fellow
- Associate consultant
- Consultant

How many years have you been in your current position?

- <5 years
- 5-10 years
- >10 years

How long you have been certified by the certification board of your primary specialty i.e. Saudi Board, American Board of Surgery, Royal College of Physicians and Surgeons of Canada, or their foreign equivalent?

- <10 years
- 10-20 years
- >20 years
- Not applicable

What is your Geographic location?

- Central region
- Northern region
- Western region
- Eastern region
- Southern region

What is the level of your trauma center?

- Level I trauma center (a sophisticated definitive care facility central to the trauma system, where all severe and complex injuries are managed by an in-house attending surgeon)
- Level II trauma center (manages severe injuries with the availability of an attending surgeon and other medical personnel)
- Level III trauma center (large community hospitals that can survey, resuscitate, and stabilize all trauma victims even if they need definitive surgery; however, complex cases beyond the available resources are transferred to higher centers)
- Undesignated

Number of trauma cases seen per year?

- <50 cases
- 50-100 cases
- >100 cases

Number of ICU beds in your facility?

- < 20 beds
- 20-60 beds
- 60 beds

What is your practice type? (Select all that apply )

- Academic Teaching Hospital
- Ministry of Health Hospital
- Private Hospital
- Government (national guard, military etc.)

Common types of trauma seen? (Select all that apply )

- Fall
- Motor vehicle accidents
- Motorcycle
- Pedestrian
- blunt
- Stab
- Intracranial injuries
- Cerebral contusion or laceration
- Brainstem or cerebellar injury
- Pelvic fracture
- Femur shaft fracture
- Tibia or fibula fracture
- Spinal cord injury
- Other (free text)

### VTE prophylaxis general questions

In your institution, decision on timing of VTE thromboprophylaxis in patients with traumatic brain injury (TBI) and Spinal Cord Injury (SCI) is made by which service?

- Intensivists
- Trauma Surgeons
- General Surgeons
- Orthopedic Trauma or Spine Surgeons
- Neurosurgeons
- Consensus between surgical, critical care, neuro services

In your institution, decision on timing of VTE thromboprophylaxis in patients conservatively managed for solid organ injury is made by which service?

- Intensivists
- Trauma Surgeons
- General Surgeons
- Consensus between surgical and critical care services
- Not applicable for my specialty

Have you seen any cases of PE and DVT in TBI, SCI, and conservatively managed solid organ injury patients who were NOT on VTE thromboprophylaxis?

- Yes
- No

If yes, how many cases?

- 1-3 cases
- 4-6 cases
- >7 cases

Have you seen any complications after starting VTE thromboprophylaxis in TBI , SCI, and conservatively managed solid organ injury ?

- Yes
- No

If Yes, how many cases?

- 1-3 cases
- 4-6 cases
- >7 cases

What types of complication? (Choose all that apply )

- Intracranial hemorrhage
- Spinal hematoma
- Retroperitoneal bleeding
- Activation of massive transfusion protocol (MTP)
- Increase blood products transfusion without the need to activate MTP
- Others (free text)

You consider the practice pattern of VTE thromboprophylaxis in TBI, SCI, and conservatively managed solid organ injury patients in your institution to be

- Conservative
- Appropriate
- Aggressive
- Undecided

Any protocol for VTE thromboprophylaxis in TBI, SCI, and conservatively managed solid organ injury patients has been implemented in your institution?

- Yes
- No
- In progress of developing one

Are you aware of any guidelines for VTE thromboprophylaxis in TBI, SCI, and conservatively managed solid organ injury patients? (Select all that apply)

- Eastern Association for the Surgery of Trauma (EAST)
- American College of Chest Physicians (ACCP)

- Brain Trauma Foundation
- Neurocritical Care Society
- None

#### Clinical Scenarios

A 53-year-old patient sustains a small subdural hematoma, and multiple fractures (femur, stable pelvis and ribs). You would start VTE thromboprophylaxis :

- In 24 h without a repeated CT head
- In 24 h if a repeated CT head is stable
- In 48 h without a repeated CT head
- In 48 h if a repeated CT head is stable
- In 72 h without a repeated CT head
- In 72 h if a repeated CT head is stable
- Not applicable to my specialty

A 24-year-old otherwise healthy male underwent a ventriculostomy for an isolated traumatic cerebral contusion. You would start VTE thromboprophylaxis

- In 24 h without a repeated CT head
- In 24 h if a repeated CT head is stable
- In 48 h without a repeated CT head
- In 48 h if a repeated CT head is stable
- In 72 h without a repeated CT head
- In 72 h if a repeated CT head is stable
- Wait until the ICP is within normal limits
- Wait until the ventriculostomy is removed
- Not applicable to my specialty

A 34-year-old male who is admitted to the neurosurgical ICU after all-terrain vehicle (ATC) crash. The patient has a spinal cord injury, and multiple rib fracture. You would start VTE thromboprophylaxis

- In 24 h without a repeated CT spine
- In 24 h if a repeated CT spine is stable
- In 48 h without a repeated CT spine
- In 48 h if a repeated CT spine is stable
- In 72 h without a repeated CT spine
- In 72 h if a repeated CT spine is stable
- Not applicable to my specialty

25-year-old male with Gun Shot Wound (GSW) in the abdomen. CT abdomen showed the trajectory of the GSW through the liver only and no other injury was identified. He is clinically stable and no evidence of active bleeding. Plan for conservative management. You would start VTE thromboprophylaxis

- In 24 h without a repeated CT abdomen
- In 24 h if a repeated CT abdomen is stable
- In 48 h without a repeated CT abdomen
- In 48 h if a repeated CT abdomen is stable
- In 72 h without a repeated CT abdomen
- In 72 h if a repeated CT abdomen is stable
- Other (please specify)
- Not applicable to my specialty

35-year-old male involved in a motor vehicle accident (MVA). CT abdomen showed grade IV splenic injury. No sign of active bleeding, he was hemodynamically stable, and you planned for conservative management. You would start VTE thromboprophylaxis

- In 24 h without a repeated CT abdomen
- In 24 h if a repeated CT abdomen is stable
- In 48 h without a repeated CT abdomen
- In 48 h if a repeated CT abdomen is stable
- In 72 h without a repeated CT abdomen
- In 72 h if a repeated CT abdomen is stable
- Not applicable to my specialty

### Factors associated with delayed initiation of VTE prophylaxis

In your opinion, what are your perceived barriers to early initiation of VTE prophylaxis within 72 h of injury (Select all that apply)

- Lack of concrete evidence
- Lack of institutional specific protocol
- Lack of communication between surgical, critical care, neuro services
- Multiple surgical interventions (laparotomy, craniotomy, ICP monitor insertions, etc.)
- Severe head injuries
- Massive early transfusion and increasing blood transfusion requirements ( >6 units of pRABC within 12 hours of injury)
- Coagulopathy and severe thrombocytopenia
- Mean ISS >25
- APACHE II score >25
- Others (free text)

In your opinion, what are the important factors in determining eligibility for early VTE initiation in trauma patients?

Short Text Box

### VTE prophylaxis preferred agents, dosing, & conclusion

What options for prevention of VTE in trauma patients most commonly used in your institution?

- Mechanical if no contraindication (e.g. lower extremity injuries)
- Pharmacological
- Combination of mechanical and pharmacological
- IVC filter

If pharmacological, which agents? Select all that apply

- Unfractionated heparin (UFH)
- Low molecular weight heparin (LMWH)
- Fondaparinux
- Warfarin

If UFH, what dose?

- 5000 units every 12 hours

- 5000 units every 8 hours
- 2500 units every 12 hours
- 2500 units every 8 hours
- Undecided

If LMWH, what dose of enoxaparin?

- 40 mg every 12 hours
- 30 mg every 12 hours
- 40 mg every 24 hours
- Weight based dosing after discussion with clinical pharmacist
- Undecided

Are you utilizing anti-Xa level for adjustment of LMWH for VTE prophylaxis in trauma patients?

- Yes
- No

Are you interested in a multicenter prospective RCT to evaluate the safety and efficacy for early (<48 h) vs. late (>48 h) pharmacological VTE prophylaxis in trauma patients?

- Yes
- No

If yes, please write down your contact information or the institution contact information (optional)
Short Text Box

**Supplementary Table 3: perceived barriers for early initiation of PVTE-Px †**

| | Total cohort N/# of responses (%) | Intensivists N/# of responses (%) | Surgeons [$] N/#of responses (%) | Other specialties* N/# of responses (%) | P value |
|---|---|---|---|---|---|
| Lack of concrete evidence | 17/102 (16.67) | 12/65 (18.46) | 5/23 (21.74) | 0/14 (0) | 0.0594 |
| Lack of institution specific protocol | 34/ 102 (33.33) | 23/ 65 (35.38) | 9/23 (39.13) | 2/14 (14.29) | 0.2119 |
| Lack of communication between services | 30/102 (29.41) | 19/65 (29.23) | 8/23 (34.78) | 3/14 (21.43) | 0.6813 |
| Multiple surgical interventions (laparotomy, craniotomy, ICP monitor insertions) | 42/102 (41.17) | 26/65 (40) | 9/23 (39.13) | 7/14 (50) | 0.7710 |
| Severe head injuries | 25/102 (24.51) | 19/65 (29.23) | 4/23 (17.39) | 2/14 (14.29) | 0.3133 |
| Massive early transfusion and increasing blood transfusion requirements (>6 units of pRBC within 12 hours of injury) | 20/102 (19.61) | 16/65 (24.62) | 3/23 (13.04) | 1/14 (7.14) | 0.1829 |
| Coagulopathy and severe thrombocytopenia | 37/102 (36.27) | 28/65 (43.08) | 7/23 (30.43) | 2/14 (14.29) | 0.0818 |
| Mean Injury severity score >25 | 7/102 (6.86) | 7/65 (10.77) | 0/23 (0) | 0/14 (0) | 0.0369 |
| APACHE II score >25 | 1/102 (0.98) | 1/ 65 (1.54) | 0/23 (0) | 0/14 (0) | 0.6355 |
| Others ( free text) [#] | 3/102 (2.94) | 1/65 (1.54) | 1/23 (4.35) | 1/14 (7.14) | 0.5210 |

**Supplementary Figure 1: Forest plots for unadjusted perceived barriers for early initiation of PVTE-Px**

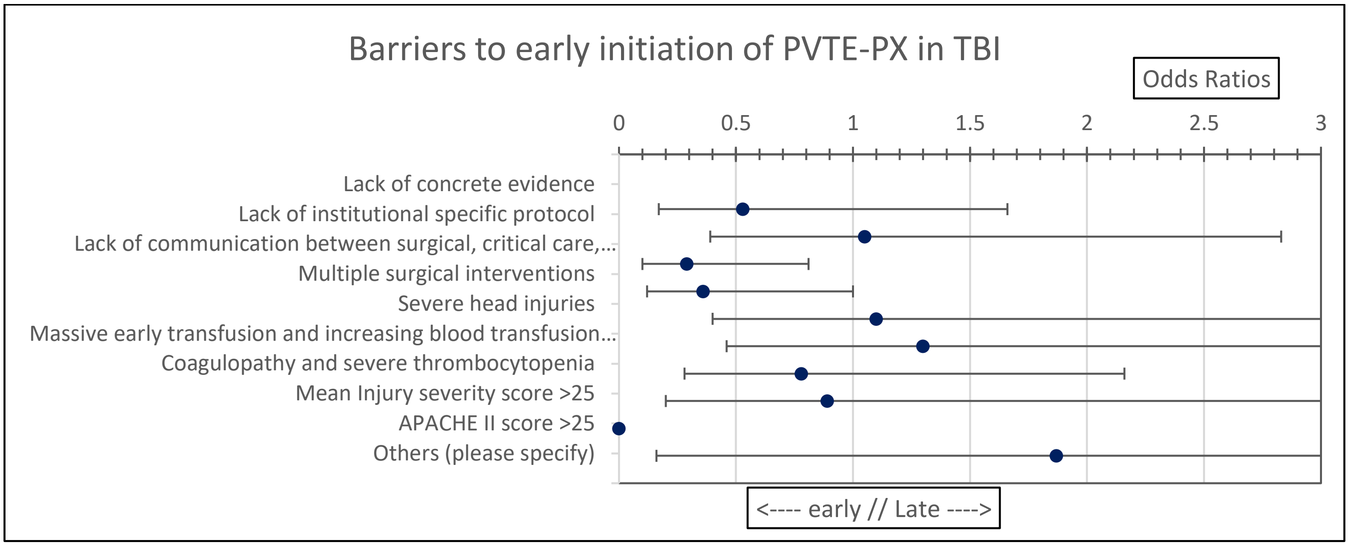


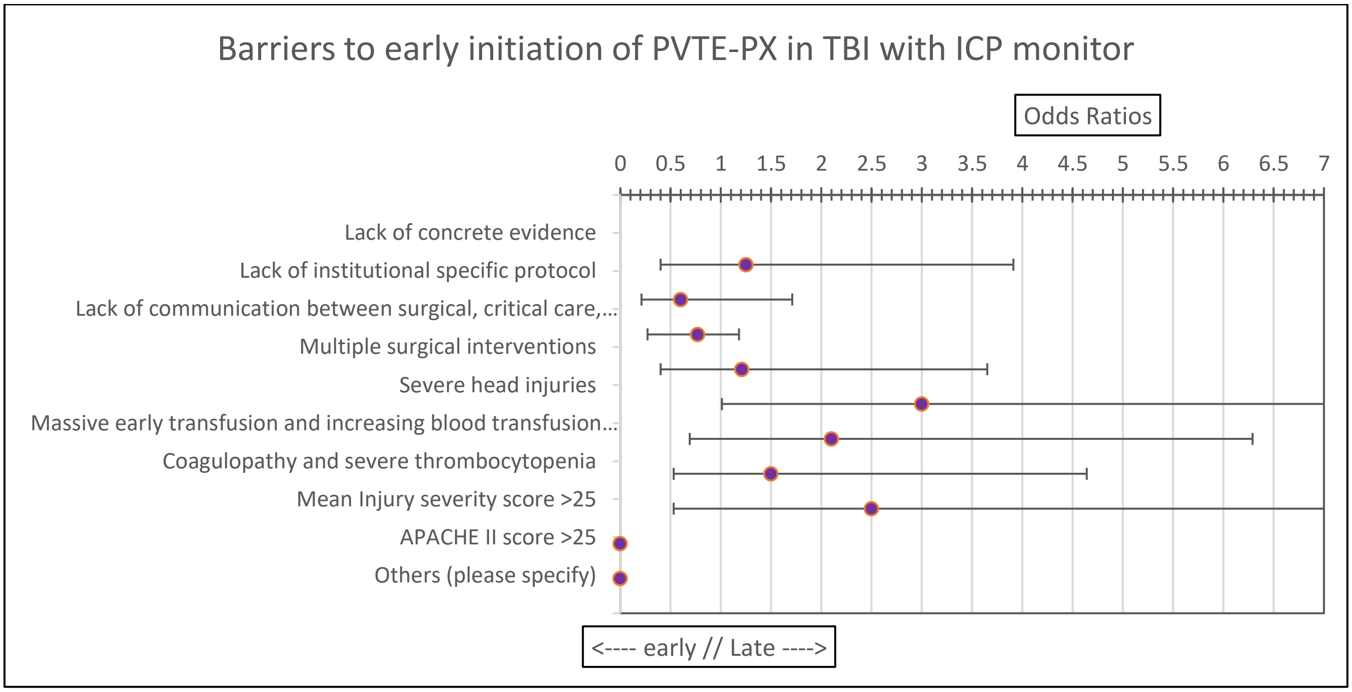

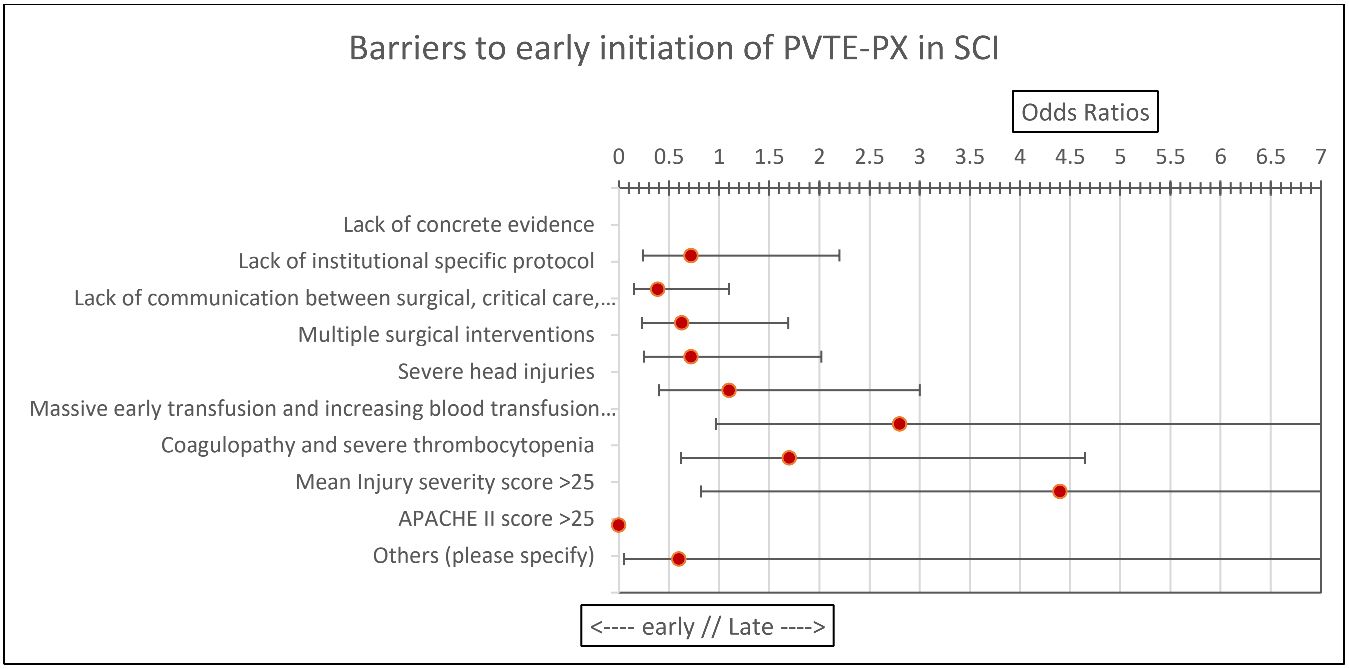
Barriers to early initiation of PVTE-PX in SCI
Odds Ratios
0 0.5 1 1.5 2 2.5 3 3.5 4 4.5 5 5.5 6 6.5 7
Lack of concrete evidence
Lack of institutional specific protocol
Lack of communication between surgical, critical care,...
Multiple surgical interventions
Severe head injuries
Massive early transfusion and increasing blood transfusion...
Coagulopathy and severe thrombocytopenia
Mean Injury severity score >25
APACHE II score >25
Others (please specify)
<---- early // Late ---->

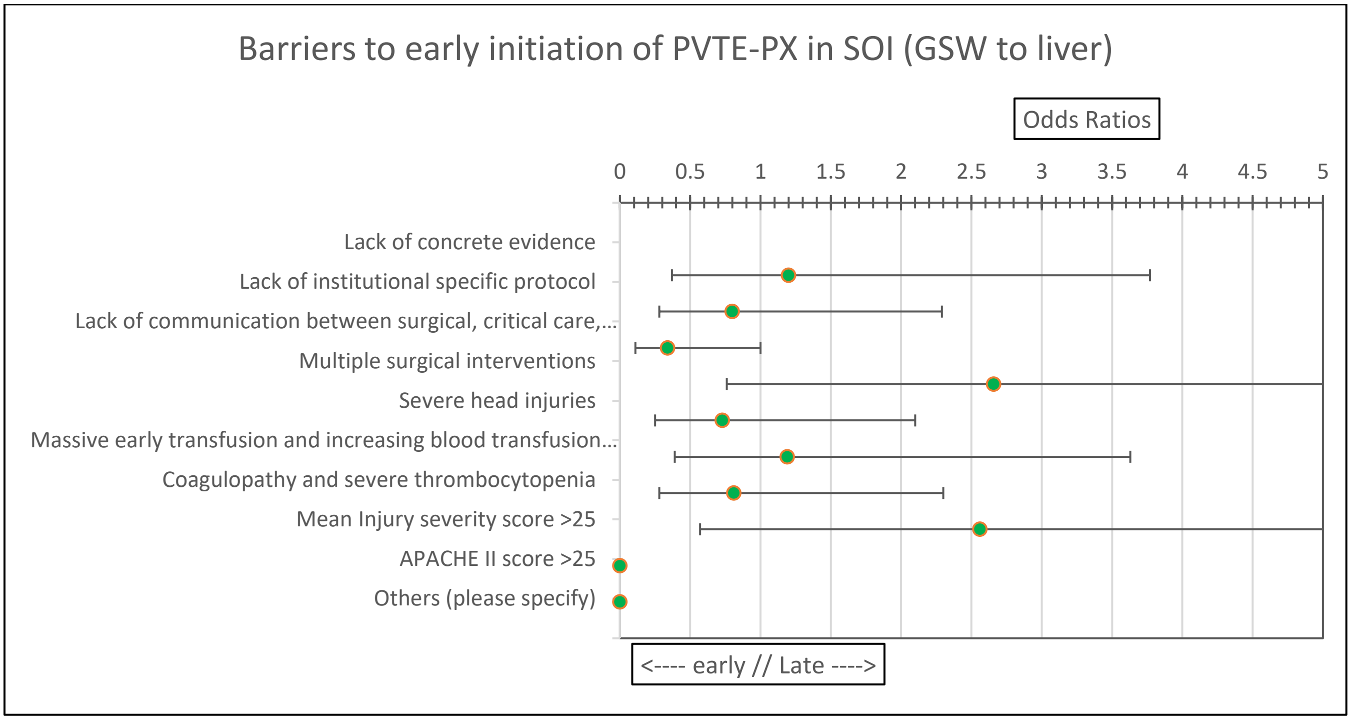
Barriers to early initiation of PVTE-PX in SOI (GSW to liver)
Odds Ratios
0 0.5 1 1.5 2 2.5 3 3.5 4 4.5 5
Lack of concrete evidence
Lack of institutional specific protocol
Lack of communication between surgical, critical care,...
Multiple surgical interventions
Severe head injuries
Massive early transfusion and increasing blood transfusion...
Coagulopathy and severe thrombocytopenia
Mean Injury severity score >25
APACHE II score >25
Others (please specify)
<---- early // Late ---->

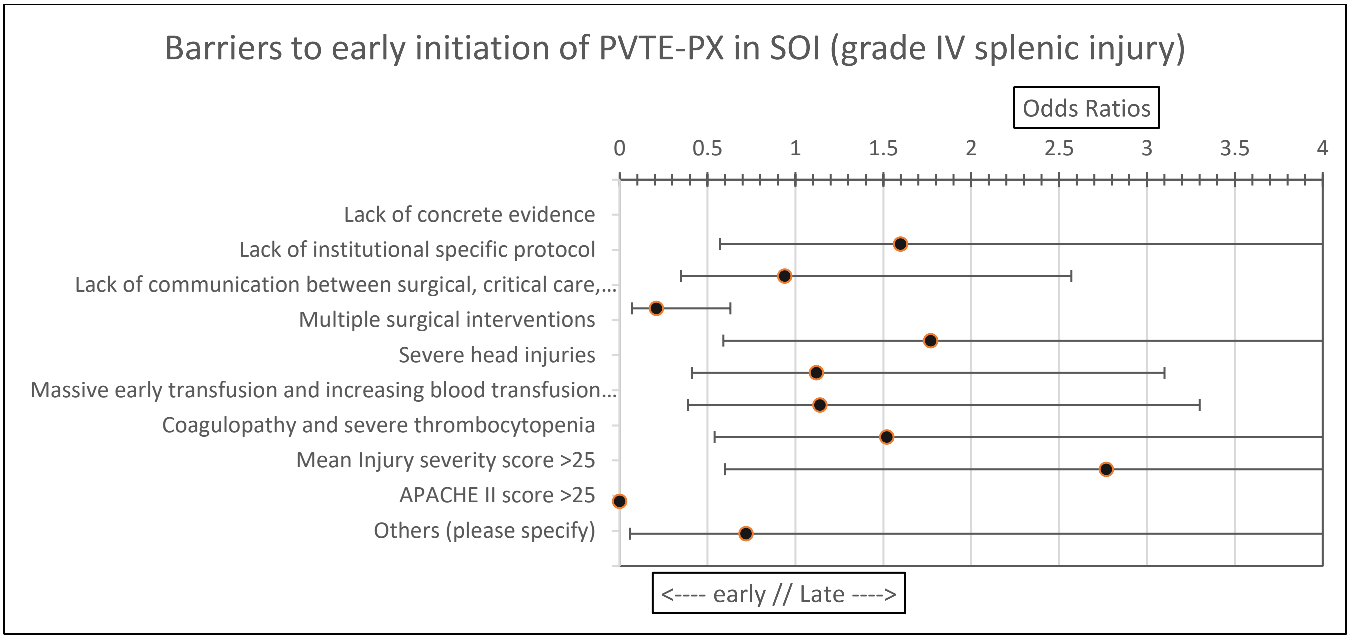
Barriers to early initiation of PVTE-PX in SOI (grade IV splenic injury)
Odds Ratios
0
0.5
1
1.5
2
2.5
3
3.5
4
Lack of concrete evidence
Lack of institutional specific protocol
Lack of communication between surgical, critical care,...
Multiple surgical interventions
Severe head injuries
Massive early transfusion and increasing blood transfusion...
Coagulopathy and severe thrombocytopenia
Mean Injury severity score >25
APACHE II score >25
Others (please specify)
<---- early // Late ---->

**Supplementary Table 4: comparison between different responses in clinical scenarios according to the positions**

| Clinical Scenarios | Total cohort N/# of responses (%) | Fellows N/# of responses (%) | Associate consultants N/# of responses (%) | Consultants N/# of responses (%) |
|---|---|---|---|---|
| **TBI** | | | | |
| In 24 h without a repeated CT head | 4/64 (6.25) | 1/14 (7.14) | 0/6 (0) | 3/44 (6.82) |
| In 24 h if a repeated CT head is stable | 26/64 (40.63) | 3/14 (21.43) | 1/6 (16.67) | 22/44 (50) |
| In 48 h without a repeated CT head | 3/64 (4.69) | 1/14 (7.14) | 0/6 (0) | 2/44 (4.55) |
| In 48 h if a repeated CT head is stable | 17/64 (26.56) | 5/14 (35.71) | 3/6 (50) | 9/44 (20.45) |
| In 72 h without a repeated CT head | 1/64 (1.56) | 0/14 (0) | 0/6 (0) | 1/44 (2.27) |
| In 72 h if a repeated CT head is stable | 11/64 (17.19) | 4/14 (28.57) | 2/6 (33.33) | 5/44 (11.36) |
| **TBI with ICP monitor** | | | | |
| In 24 h without a repeated CT head | 7/64 (10.94) | 2/14 (14.29) | 0/6 (0) | 5/44 (11.36) |
| In 24 h if a repeated CT head is stable | 26/64 (40.63) | 5/14 (35.71) | 2/6 (3.33) | 19/44 (43.18) |
| In 48 h without a repeated CT head | 5/64 (7.81) | 1/14 (7.14) | 1/6 (16.67) | 3/44 (6.82) |
| In 48 h if a repeated CT head is stable | 10/64 (15.63) | 3/14 (21.43) | 0/6 (0) | 7/44 (15.91) |
| In 72 h without a repeated CT head | 7/64 (10.94) | 2/14 (14.29) | 3/6 (50) | 2/44 (4.55) |
| In 72 h if a repeated CT head is stable | 3/64 (4.69) | 0/14 (0) | 0/6 (0) | 3/44 (6.82) |
| Wait until the ICP is within normal | 1/64 (1.56) | 0/14 (0) | 0/6 (0) | 1/44 (2.27) |
| Wait until the ventriculostomy is removed | 5/64 (7.81) | 1/14 (7.14) | 0/6 (0) | 4/44 (9.09) |
| **SCI** | | | | |
| In 24 h without a repeated CT spine | 21/64 (32.81) | 4/14 (28.57) | 0/6 (0) | 17/44 (38.64) |
| In 24 h if a repeated CT spine is stable | 15/64 (23.43) | 3/14 (21.43) | 1/6 (16.67) | 11/44 (25) |
| In 48 h without a repeated CT spine | 3/64 (4.69) | 0/14 (0) | 1/6 (16.67) | 2/44 (4.55) |
| In 48 h if a repeated CT spine is stable | 12/64 (18.75) | 6/14 (42.86) | 2/6 (33.33) | 4/44 (9.09) |
| In 72 h without a repeated CT spine | 2/64 (3.13) | 1/14 (7.14) | 1/6 (16.67) | 0/44 (0) |
| In 72 h if a repeated CT spine is stable | 4/64 (6.25) | 0/14 (0) | 1/6 (16.67) | 3/44 (6.82) |
| **NOR solid organ injuries: Gun Shot Wound (GSW) through the Liver** | | | | |
| In 24 h without a repeated CT abdomen | 13/64 (20.31) | 2/14 (14.29) | 1/6 (16.67) | 10/44 (22.73) |
| In 24 h if a repeated CT abdomen is stable | 10/64 (15.63) | 3/14 (21.43) | 2/6 (33.33) | 5/44 (11.36) |
| In 48 h without a repeated CT abdomen | 15/64 (23.44) | 5/14 (35.71) | 0/6 (0) | 10/44 (22.73) |
| In 48 h if a repeated CT abdomen is stable | 7/64 (10.94) | 1/14 (7.14) | 2/6 (33.33) | 4/44 (9.09) |
| In 72 h without a repeated CT abdomen | 4/64 (6.25) | 0/14 (0) | 0/6 (0) | 4/44 (9.09) |
| In 72 h if a repeated CT abdomen is stable | 7/64 (10.94) | 2/14 (12.29) | 1/6 (16.67) | 4/44 (9.09) |
| **NOR solid organ injuries: grade IV splenic injury** | | | | |
| In 24 h without a repeated CT abdomen | 9/63 (14.29) | 2/14 (14.29) | 3/6 (50) | 4/43 (9.30) |
| In 24 h if a repeated CT abdomen is stable | 10/63 (15.87) | 3/14 (21.43) | 1/6 (16.67) | 6/43 (13.95) |
| In 48 h without a repeated CT abdomen | 11/63 (17.46) | 0/14 (0) | 1/6 (16.67) | 10/43 (23.26) |
| In 48 h if a repeated CT abdomen is stable | 8/63 (12.70) | 2/14 (14.29) | 1/6 (16.67) | 5/43 (11.63) |
| In 72 h without a repeated CT abdomen | 5/63 (7.94) | 1/14 (7.14) | 0/6 (0) | 4/43 (9.30) |
| In 72 h if a repeated CT abdomen is stable | 13/63 (20.63) | 3/14 (21.43) | 0/6 (0) | 10/43 (23.36) |

**Supplementary Table 5: comparison of different responses in clinical scenarios according to practicing years**

| Clinical Scenarios | Total cohort N/# of responses (%) | < 5 years N/# of responses (%) | 5-10 years N/# of responses (%) | >10 years N/# of responses (%) |
|---|---|---|---|---|
| **TBI** | | | | |
| In 24 h without a repeated CT head | 4/65 (6.15) | 1/27 (3.7) | 2/13 (15.38) | 1/25 (4) |
| In 24 h if a repeated CT head is stable | 26/65 (40) | 10/27 (37.04) | 6/13 (46.15) | 10/25 (40) |
| In 48 h without a repeated CT head | 3/65 (4.62) | 1/27 (3.7) | 0/13 (0) | 2/25 (8) |
| In 48 h if a repeated CT head is stable | 17/65 (26.15) | 7/27 (25.93) | 4/13 (30.77) | 6/25 (24) |
| In 72 h without a repeated CT head | 1/65 (1.54) | 0/27 (0) | 0/13 (0) | 1/25 (4) |
| In 72 h if a repeated CT head is stable | 12/65 (18.46) | 8/27 (29.63) | 1/13 (7.69) | 3/25 (12) |
| **TBI with ICP monitor** | | | | |
| In 24 h without a repeated CT head | 7/65 (17.77) | 2/27 (7.41) | 1/13 (7.69) | 4/25 (16) |
| In 24 h if a repeated CT head is stable | 26/65 (40) | 12/27 (44.44) | 9/13 (69.23) | 5/25 (20) |
| In 48 h without a repeated CT head | 5/65 (7.70) | 2/27 (7.41) | 0/13 (0) | 3/25 (12) |
| In 48 h if a repeated CT head is stable | 10/65 (15.38) | 3/27 (11.11) | 2/13 (15.38) | 5/25 (20) |
| In 72 h without a repeated CT head | 7/65 (17.77) | 2/27 (7.41) | 1/13 (7.69) | 4/25 (16) |
| In 72 h if a repeated CT head is stable | 3/65 (4.62) | 1/27 (3.7) | 0/13 (0) | 2/25 (8) |
| Wait until the ICP is within normal | 1/65 (1.54) | 1/27 (3.7) | 0/13 (0) | 0/25 (0) |
| Wait until the ventriculostomy is removed | 6/65 (9.23) | 4/27 (14.81) | 0/13 (0) | 2/25 (8) |
| **SCI** | | | | |
| In 24 h without a repeated CT spine | 21/65 (32.31) | 10/27 (37.04) | 5/13 (38.46) | 6/25 (24) |
| In 24 h if a repeated CT spine is stable | 15/65 (23.07) | 7/27 (25.93) | 3/13 (23.08) | 5/25 (20) |
| In 48 h without a repeated CT spine | 3/65 (4.62) | 1/27 (3.70) | 0/13 (0) | 2/25 (8) |
| In 48 h if a repeated CT spine is stable | 13/65 (20) | 6/27 (22.22) | 1/13 (7.69) | 6/25 (24) |
| In 72 h without a repeated CT spine | 2/65 (3.08) | 0/27 (0) | 2/13 (15.38) | 0/25 (0) |
| In 72 h if a repeated CT spine is stable | 4/65 (6.15) | 0/27 (0) | 1/13 (7.69) | 3/25 (12) |
| **NOR solid organ injuries: Gun Shot Wound (GSW) through the Liver** | | | | |
| In 24 h without a repeated CT abdomen | 13/65 (20) | 7/27 (25.93) | 3/13 (23.08) | 3/25 (12) |
| In 24 h if a repeated CT abdomen is stable | 10/65 (15.38) | 4/27 (14.81) | 4/13 (30.77) | 2/25 (8) |
| In 48 h without a repeated CT abdomen | 15/65 (23.07) | 8/27 (29.63) | 2/13 (15.38) | 5/25 (20) |
| In 48 h if a repeated CT abdomen is stable | 7/65 (17.77) | 3/27 (11.11) | 2/13 (15.38) | 2/25 (8) |
| In 72 h without a repeated CT abdomen | 4/65 (6.15) | 0/27 (0) | 1/13 (7.69) | 3/25 (12) |
| In 72 h if a repeated CT abdomen is stable | 8/65 (12.31) | 3/27 (11.11) | 0/13 (0) | 5/25 (20) |
| **NOR solid organ injuries : grade IV splenic injury** | | | | |
| In 24 h without a repeated CT abdomen | 9/64 (14.06) | 4/27 (14.81) | 3/13 (23.08) | 2/24 (8.33) |
| In 24 h if a repeated CT abdomen is stable | 10/64 (15.63) | 4/27 (14.81) | 5/13 (38.46) | 1/24 (4.17) |
| In 48 h without a repeated CT abdomen | 11/64 (17.18) | 5/27 (18.52) | 2/13 (15.38) | 4/24 (16.67) |
| In 48 h if a repeated CT abdomen is stable | 8/64 (12.5) | 3/27 (11.11) | 1/13 (7.69) | 4/24 (16.67) |
| In 72 h without a repeated CT abdomen | 5/64 (7.81) | 2/27 (7.41) | 1/13 (7.69) | 2/24 (8.33) |
| In 72 h if a repeated CT abdomen is stable | 13/64 (20.31) | 4/27 (14.81) | 0/13 (0) | 9/24 (37.50) |

**Supplementary Table 6: comparison of different responses in clinical scenarios according to the board certification**

| Clinical Scenarios | Total cohort N/# of responses (%) | board certified <10 N/# of responses (%) | board certified 10-20 N/# of responses (%) | board certified >20 N/# of responses (%) |
|---|---|---|---|---|
| **TBI** | | | | |
| In 24 h without a repeated CT head | 4/61 (6.56) | 1/22 (4.55) | 3/32 (9.38) | 0/7 (0) |
| In 24 h if a repeated CT head is stable | 26/61 (42.62) | 9/22 (40.91) | 13/32 (40.63) | 4/7 (57.14) |
| In 48 h without a repeated CT head | 2/61 (3.28) | 0/22 (0) | 2/32 (6.25) | 0/7 (0) |
| In 48 h if a repeated CT head is stable | 16/61 (26.23) | 9/22 (40.91) | 5/32 (15.63) | 2/7 (28.57) |
| In 72 h without a repeated CT head | 1/61 (1.64) | 0/22 (0) | 1/32 (3.13) | 0/7 (0) |
| In 72 h if a repeated CT head is stable | 10/61 (16.39) | 3/22 (13.64) | 6/32 (18.75) | 1/7 (14.29) |
| **TBI with ICP monitor** | | | | |
| In 24 h without a repeated CT head | 7/61 (11.48) | 2/22 (9.09) | 4/32 (12.5) | 1/7 (14.29) |
| In 24 h if a repeated CT head is stable | 26/61 (42.62) | 12/22 (54.55) | 11/32 (34.38) | 3/7 (42.86) |
| In 48 h without a repeated CT head | 5/61 (8.19) | 0/22 (0) | 4/32 (12.50) | 1/7 (14.29) |
| In 48 h if a repeated CT head is stable | 8/61 (13.11) | 2/22 (9.09) | 5/32 (15.63) | 1/7 (14.29) |
| In 72 h without a repeated CT head | 6/61 (9.84) | 2/22 (9.09) | 3/32 (9.38) | 1/7 (14.29) |
| In 72 h if a repeated CT head is stable | 3/61 (4.91) | 0/22 (0) | 3/32 (9.38) | 0/7 (0) |
| Wait until the ICP is within normal | 1/61 (1.64) | 1/22 (4.55) | 0/32 (0) | 0/7 (0) |
| Wait until the ventriculostomy is removed | 5/61 (8.19) | 3/22 (13.64) | 2/32 (6.25) | 0/7 (0) |
| **SCI** | | | | |
| In 24 h without a repeated CT spine | 21/61 (34.43) | 10/22 (45.45) | 10/32 (31.25) | 1/7 (14.29) |
| In 24 h if a repeated CT spine is stable | 14/61 (22.95) | 5/22 (22.73) | 7/32 (21.88) | 2/7 (28.57) |
| In 48 h without a repeated CT spine | 3/61 (4.91) | 0/22 (0) | 2/32 (6.25) | 1/7 (14.29) |
| In 48 h if a repeated CT spine is stable | 10/61 (16.39) | 3/22 (13.64) | 5/32 (15.63) | 2/7 (28.57) |
| In 72 h without a repeated CT spine | 2/61 (3.28) | 2/22 (9.09) | 0/32 (0) | 0/7 (0) |
| In 72 h if a repeated CT spine is stable | 4/61 (6.56) | 0/22 (0) | 3/32 (9.38) | 1/7 (14.29) |
| **NOR solid organ injuries: Gun Shot Wound (GSW) through the Liver** | | | | |
| In 24 h without a repeated CT abdomen | 13/61 (21.31) | 6/22 (27.27) | 7/32 (21.88) | 0/ 7 (0) |
| In 24 h if a repeated CT abdomen is stable | 9/61 (14.75) | 4/22 (18.18) | 3/32 (9.38) | 2/ 7 (28.57) |
| In 48 h without a repeated CT abdomen | 15/61 (24.59) | 7/22 (31.82) | 6/32 (18.75) | 2/ 7 (28.57) |
| In 48 h if a repeated CT abdomen is stable | 7/61 (11.48) | 4/22 (18.18) | 2/32 (6.25) | 1/ 7 (14.29) |
| In 72 h without a repeated CT abdomen | 4/61 (6.56) | 0/22 (0) | 4/32 (12.5) | 0/ 7 (0) |
| In 72 h if a repeated CT abdomen is stable | 6/61 (9.83) | 0/22 (0) | 5/32 (15.63) | 1/ 7 (14.29) |
| **NOR solid organ injuries: grade IV splenic injury** | | | | |
| In 24 h without a repeated CT abdomen | 9/60 (15) | 4/22 (18.18) | 4/31 (12.9) | 1/7 (14.29) |
| In 24 h if a repeated CT abdomen is stable | 9/60 (15) | 3/22 (13.64) | 6/31 (19.35) | 0/7 (0) |
| In 48 h without a repeated CT abdomen | 11/60 (18.33) | 3/22 (13.64) | 6/31 (19.35) | 2/7 (28.57) |
| In 48 h if a repeated CT abdomen is stable | 8/60 (13.33) | 3/22 (13.64) | 3/31 (9.68) | 2/7 (28.57) |
| In 72 h without a repeated CT abdomen | 5/60 (8.33) | 3/22 (13.64) | 2/31 (6.45) | 0/7 (0) |
| In 72 h if a repeated CT abdomen is stable | 12/60 (20) | 2/22 (9.09) | 8/31 (25.81) | 2/7 (28.57) |
